# Evaluation of AI-Generated Synthetic Data for Clinical Research in Secondary Cardiovascular Prevention among Dyslipidemia Patients

**DOI:** 10.64898/2026.06.12.26355456

**Authors:** Alice Bonomi, Pablo Josè Werba, Sebastiano Saccani, Lorenzo Li Lu, Andrea Coser, Matteo Franchi, Carola Valsecchi, Elisa Teruzzi, Alessandra Terragni, Chiara Centenaro, Marco Scatigna, Giulio Pompilio

## Abstract

**Background:** Access to high-quality clinical data is essential for advancing medical research and developing effective medical statistical and Artificial Intelligence models. However, privacy regulations and logistical barriers often hinder timely access to real-world data. Synthetic data offer a promising solution, preserving the statistical characteristics of original datasets while protecting patient privacy.

**Objectives:** This study investigates the use of synthetic data for secondary cardiovascular prevention in patients with dyslipidemia, using two real-world datasets from Centro Cardiologico Monzino.

**Methods:** Given the high dimensionality and limited sample size of the datasets, we employed a custom generative framework based on Large Language Models (LLMs). Pre-trained LLMs were fine-tuned on original clinical records to synthesize tabular data replicating source-data distributions. Fine-tuning was performed within the Centro Cardiologico Monzino’s secure infrastructure to ensure data sovereignty. We evaluate clinical utility and privacy using fidelity and privacy metrics, identifying the optimal generative model and benchmarking against traditional anonymization methods.

**Results:** Synthetic data achieved a superior trade-off than classically anonymized datasets. Real and synthetic datasets showed strong agreement, with significant distributional differences limited to few variables. Models trained on synthetic data replicated key associations from the original dataset, including therapy modification and creatine phosphokinase as predictors of SAMS, and pharmacological intensity as the main driver of LDL-C reduction.

**Conclusions:** Results support the feasibility of using synthetic data as a proxy for real-world datasets in exploratory analyses and model development. Despite slight attenuation of some effect sizes, preserved clinical relationships reinforce the validity of synthetic data in medical research.

## Introduction

Data availability plays a crucial role in medical research. Timely access to high-quality data is paramount not only in the development of reliable medical models^1^, but also in facilitating efficient patient care^2^. The rise of modern Artificial Intelligence (AI) models has fueled an even greater demand for data, since machine learning depends on vast amounts of information for development^3^.

Unfortunately, data access is hindered in practice by a number of obstacles. These include legal and ethical concerns around patient privacy^4^. Regulations like the General Data Protection Regulation (GDPR) and the Health Insurance Portability and Accountability Act (HIPAA) contribute to prolonged and costly research efforts, primarily due to the complex procedures involved in handling data access requests^5^. In addition to privacy concerns, logistical challenges further impede efficient access to high-quality data. Real-world data (RWD) may be incomplete^6–7^, error-prone^8–9^ and biased^10^.

Synthetic data generation has recently gained attention as a key technology in overcoming obstacles to medical data access^11–14^. Synthetic data are computer-generated data designed to replicate the properties and analytical value of RWD while removing sensitive information^12,15^. The substitution of RWD by synthetic data can therefore facilitate more efficient statistical analyses and the development of AI models, whilst protecting patient privacy. To achieve this, however, synthetic data must meet strict requirements for both privacy and utility^16–19^. Recent studies have shown that, when properly generated, synthetic data can satisfy these expectations across diverse applications. These applications can be broadly categorized into (i) disease-specific domains, including hepatology^20^, oncology^21–22^ and hematological malignancies^23–24^, and cardiovascular diseases^25^; and (ii) methodological domains, such as epidemiological investigations^26^, observational research, and the design and conduct of clinical trials^27–29^.

The purpose of this study is to validate the use of synthetic data in providing an accurate replicate of real medical data. Specifically, we compared RWD and synthetic data generated by Aindo’s technology^30–32^ in the context of secondary cardiovascular prevention of patients with dyslipidemia. We find that analyses of synthetic data lead to results comparable to the same analyses applied to RWD. This indicates that models trained on synthetic data can be transferred effectively to RWD applications, supporting wider use of synthetic datasets in clinical and epidemiological studies.

### Novel contributions

This paper explores the synthesis of clinical data in the context of secondary cardiovascular prevention for patients with dyslipidemia, providing the research community with access to the resulting synthetic cohorts. The source datasets present a significant methodological challenge: they feature a high-dimensional feature space (∼60 variables) coupled with a relatively modest sample size (<1000 patients). Given that the complexity of the underlying probability distribution grows exponentially with the number of variables, traditional synthetic data generation techniques often struggle to achieve satisfactory results with limited data.

To address this, we implement a generative strategy that fine-tunes pre-trained Large Language Models (LLMs), thereby leveraging both the latent clinical knowledge acquired during pre-training and the empirical distributions of the real-world datasets. By incorporating pre-trained clinical knowledge, this strategy aims to mitigate the challenges of data scarcity, which often constrain the statistical fidelity of standard synthesis models. Our methodology diverges from existing literature — which typically relies either on non-pretrained models trained from scratch^26,33^ or on zero-shot generation without domain-specific fine-tuning^34^ — by explicitly combining pre-trained medical context with fine-tuning on real-world evidence. Finally, we employ a dual-objective selection framework, utilizing both privacy and broad utility (fidelity) metrics to identify the optimal generative model.

## Methods

### Data

In this study, two different datasets of RWD regarding the ambulatory care of patients with dyslipidemia in secondary cardiovascular prevention, collected at the Centro Cardiologico Monzino, were used.

- **SAMS dataset:** This database was developed to allow the analysis of clinical factors associated with Statin-Associated Muscle Symptoms (SAMS). It includes data from 1000 patients, with 47 categorical variables and 25 numerical variables.
- **LDL-C dataset:** This dataset was developed to analyze the relationship between percent changes in LDL-C levels and the achievement rates of therapeutic goals, based on the intensity of therapeutic management, the number of treatment modifications, and the type of final therapy (monotherapy vs combination therapy, including or excluding PCSK9 inhibitors). It includes data from 700 patients, with 38 categorical variables and 18 numerical variables.

### Synthetic data generation

To generate the synthetic patient cohorts, we employ a custom synthetic data framework centered on a LLM-based generative module. The pipeline utilizes open weight pre-trained LLMs sourced from the Hugging Face repository^35^, which are subsequently fine-tuned on the real-world clinical datasets to specialize them for synthetic tabular data generation. The fine-tuning process is executed on a secure cloud-based virtual machine residing entirely within the Centro Cardiologico Monzino’s private infrastructure, ensuring that the original patient records never exit the protected environment. Through this fine-tuning process, the model learns the underlying probability distributions, multi-variable correlations, and complex structural patterns inherent in the original data. This enables the generation of synthetic records that faithfully reflect these clinical characteristics. To ensure the output strictly maintains structural integrity relative to the original schemas, we implement constrained (structured) generation techniques.

To select the optimal number of fine-tuning steps, we evaluate synthetic datasets produced at multiple checkpoints throughout the fine-tuning process. To ensure the versatility of the synthetic data, we adopt an analysis-agnostic approach for model selection that remains independent of specific downstream tasks. Each generative checkpoint is evaluated through a dual-lens framework of fidelity (broad utility) and privacy preservation. This comprehensive assessment allows us to identify a configuration that achieves a robust trade-off between high data utility and privacy protection.

We provide all the details of the synthetic data generation pipeline in the supplementary materials, including data preprocessing, structured generation and fine-tuning. Moreover, to ensure reproducibility and transparency in our findings, the complete codebase for data generation, along with the evaluation frameworks for utility and privacy, has been made available in a public repository^36^.

### Model selection

As previously outlined, to identify the optimal generative configuration, we evaluate both fidelity and privacy metrics at each evaluation checkpoint. We observe a consistent inverse relationship during extended training: as the fine-tuning duration increases, the synthetic datasets typically exhibit enhanced fidelity at the expense of reduced privacy guarantees. Consequently, we select the specific checkpoint that demonstrates the highest broad utility while strictly adhering to predefined privacy thresholds.

To contextualize the performance of our generative approach, we benchmark the synthetic data against traditional anonymization techniques. We compare our fidelity and privacy assessments to those of a suite of anonymized datasets, which are generated by injecting varying degrees of stochastic noise into the source data. This noise injection is governed by a parameter α, allowing us to evaluate the efficiency of LLM-based synthetic data generation relative to classical anonymization across a spectrum of privacy-utility trade-offs.

More details about the anonymization techniques used are presented in the Supplementary Materials.

### Fidelity assessment

Utility metrics for synthetic data are generally categorized into two distinct classes^16,37–38^:

- **Broad utility (fidelity):** This category evaluates the overall statistical alignment between the real and synthetic datasets. It includes univariate and bivariate distributions, as well as metrics that capture deeper structural patterns, such as multivariate density similarities, clustering similarities, and distinguishability.
- **Narrow utility:** These metrics compare the results of specific downstream tasks on the real and synthetic data. This evaluation ranges from the comparison of targeted descriptive statistics to benchmarking the predictive performance of Machine Learning (ML) models trained on synthetic versus original data (e.g., Train on Synthetic, Test on Real).

In the subsequent sections, we present a statistical analysis specifically designed to evaluate the clinical performance of the synthetic data relative to the source data; this analysis constitutes a narrow utility assessment. Conversely, for the purpose of efficient model selection, we focus primarily on broad utility metrics. By selecting models based on their ability to replicate the global data distribution rather than a single clinical endpoint, we ensure the resulting synthetic datasets are analysis-agnostic and maintain the versatility required for diverse research applications^39–40^.

To quantify fidelity, we employ a distinguishability metric implemented through an XGBoost discriminator. The model is tasked with performing a binary classification to differentiate between real and synthetic records, labeled as 0 and 1, respectively. The performance of this discriminator is measured using the Area Under the Curve of the Receiver Operating Characteristic (AUC-ROC). More details are provided in the Supplementary Materials.

### Privacy assessment

Quantifying residual privacy risk in synthetic data remains an active area of research. While numerous metrics exist, no single gold standard has emerged; each possesses distinct advantages and inherent limitations. In our evaluation, we employ a multi-layered approach combining distance-based metrics and empirical privacy auditing.

#### DCR-based metrics and the Privacy Score

A foundational class of privacy metrics relies on the Distance to Closest Record (DCR). While various definitions of DCR exist in the literature, they all fundamentally rely on calculating the proximity of each synthetic record to its nearest neighbor in the real training set. A critically low DCR for certain synthetic records suggests that the generative model may be partially “memorizing” the source data, often referred to as *overfitting*. To confirm this, the DCR is often compared against a baseline DCR calculated using a holdout (test) set not used during training. If the DCR relative to the training data is statistically significantly smaller than the DCR relative to the test data, it indicates a propensity for overfitting.

Computing any DCR-based metric requires a robust distance function, which is built upon two fundamental components:

1. **Encoding:** A function mapping each record into a specific numerical space.
2. **Distance computation:** A measure of proximity between encoded records.

For our evaluation, we implement and apply a DCR-based metric, that we call the *Privacy Score p*. The details about the computation of the *Privacy Score* are presented in the Supplementary Materials.

#### Attack-based metrics and Anonymeter

While DCR-like metrics are computationally efficient and widely adopted by practitioners, research has demonstrated that they can fail to detect meaningful privacy leaks in certain case^41^. Consequently, in recent years adversarial attack frameworks have gained prominence^12,42–45^. In these scenarios, an attacker utilizes the published synthetic dataset alongside auxiliary information to gain knowledge about the original training data. The most common of these are Membership Inference Attacks (MIAs)—where an attacker attempts to determine if a known target record was used in training—and Attribute Inference Attacks (AIAs)—where the attacker has access to a partial training record and tries to infer knowledge about the remaining attributes.

Unfortunately, the practical implementation of privacy metrics based on MIAs is often hindered by unrealistic requirements for auxiliary information or prohibitive computational costs. In our specific context, these factors render the calculation of standard MIAs unfeasible. Many such attacks require an adversary to possess a large reference set—a collection of records following the same statistical distribution as the training data. Given the inherent scarcity of our original clinical datasets, extracting such a reference set without compromising the integrity of the study is not possible.

Furthermore, state-of-the-art MIAs frequently necessitate the training of a large ensemble of “shadow models” to simulate the behavior of the target generator. This requirement is computationally incompatible with our framework, as the memory overhead and time required for each individual LLM fine-tuning session make large-scale model replication impractical. Consequently, we prioritize more efficient yet robust auditing methods that align with the resource constraints.

Although state-of-the-art MIAs remain computationally inaccessible, we employ an attack-based metric using the publicly available Anonymeter tool^46^. Anonymeter provides a unified framework for quantifying risks associated with singling out, linkability, and attribute inference. The tool simulates adversarial behavior through a series of automatically generated “guesses” derived directly from the synthetic dataset. The residual privacy risk is quantified by comparing the success rate of these guesses on the training set against a designated control set. This comparison is essential for distinguishing between a genuine privacy breach—where the model has memorized specific training instances—and the replication of global statistical patterns, which constitutes the primary utility of synthetic data. This distinction is particularly critical in the context of AIAs, where a model may correctly “guess” an attribute simply by having accurately learned the underlying clinical correlations common to the entire patient population.

Further technical details regarding the Anonymeter evaluation—including our choice of attack vectors and their corresponding parameters—are documented in the Supplementary Materials.

### Checkpoint selection

To identify the optimal generator, model checkpoints are saved at regular intervals during the fine-tuning process. We then generate synthetic datasets for each candidate checkpoint and calculate the previously defined fidelity and privacy metrics. To facilitate a comparative assessment, the results are visualized using two-dimensional utility-privacy plots, with the classical anonymization benchmarks included as a performance baseline.

Our selection framework incorporates a single fidelity metric—the XGBoost discriminator AUC-ROC—and three privacy metrics: the Privacy Score, the Anonymeter singling out attack (utilizing multivariate predicates), and the Anonymeter linkability attack.

We exclude the Anonymeter attribute inference attack from the formal selection criteria, as the results exhibit high variance when applied to LLM-generated synthetic data. However, extensive testing across multiple “secret” target columns confirms that the adversarial success rate for the synthetic data remains consistently lower than that of the anonymized benchmarks. While we report specific inference attack results using clinical target variables as secret columns for illustrative purposes, these are not used to determine the final model checkpoint.

The three fidelity-versus-privacy trade-off plots allow for a qualitative Pareto-optimal selection, identifying the generative model that provides the most robust balance between statistical utility and privacy preservation.

### Statistical analysis

For continuous variables, differences in the frequency distribution between real and synthetic datasets were assessed using the non-parametric Wilcoxon rbnchank-sum test. For categorical variables, distributional similarity was evaluated using the Jensen-Shannon Divergence (JSD).

Missing data were imputed using a multiple imputation technique, using the PROC MI procedure in SAS software (version 9.4; SAS Institute Inc., Cary, NC, USA).

To investigate the association between clinical factors and SAMS, a logistic regression model with stepwise selection and cross-validation was employed. The datasets were randomly split into a training set (50%) and a validation set (50%). Stepwise selection was performed on the training set, and the model’s performance was subsequently evaluated on the validation set. This procedure was repeated over 200 iterations. Variables were retained if both their inclusion and confirmation rates exceeded 70%.

Following variable selection, a logistic regression model was fitted using the selected predictors in both the real and synthetic datasets. Odds ratios (ORs) and their corresponding 95% CI were compared using the CI overlap metric, calculated as follows:

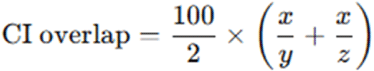

where *x* is the width of the overlapping portion of the two CIs, *y* is the width of the CI from the original dataset, and *z* is the width of the CI from the synthetic dataset.

In the second case study, a General Linear Model (GLM) was implemented to evaluate the relationship between changes in LDL-C levels, clinical factors, and therapeutic management, including treatment intensity and therapy modifications. Stepwise selection and cross-validation were also applied. Results were reported as beta coefficients and standard errors. A bootstrap procedure with 5,000 iterations was conducted in both the synthetic and original datasets to ensure robustness of the estimates.

### Ethical Approval

The research project (CCM-NP1108) was evaluated and authorized by an independent body, the “CCM Retrospective Studies Data Governance Board,” considering its scientific relevance, its relevance to the research areas of the Monzino Cardiology Center, and the risk-benefit ratio from a scientific, ethical, and personal data protection standpoint.

Our study adhered to the Declaration of Helsinki.

## Results

### Checkpoint selection

As established in the methodological framework, we identify the optimal generative model by manually assessing the privacy-utility trade-off throughout the fine-tuning process. We employ the XGBoost discriminator AUC-ROC as the fidelity metric (where lower values indicate superior fidelity) and evaluate privacy risk through three distinct lenses:

- **Privacy Score:** Visualized in Figure 1 (SAMS dataset) and Figure 2 (LDL-C dataset). We plot 1 − *p* to provide a direct measure of privacy risk. We use *k* = 1 and *q* = 0. 1.
- **Singling-out attack:** Evaluated via Anonymeter using multivariate predicates and 500 attacks. The results are in Figure 3 and Figure 4.
- **Linkability attack:** Evaluated via Anonymeter by using *k* = 1 and randomly selecting 20 attributes to define the bifurcated subspaces. Due to the constraints of the holdout set size, the number of attack iterations is set to 100 for the SAMS dataset and 70 for the LDL-C dataset. The results are visualized in Figure 5 and Figure 6.

**Figure 1.**
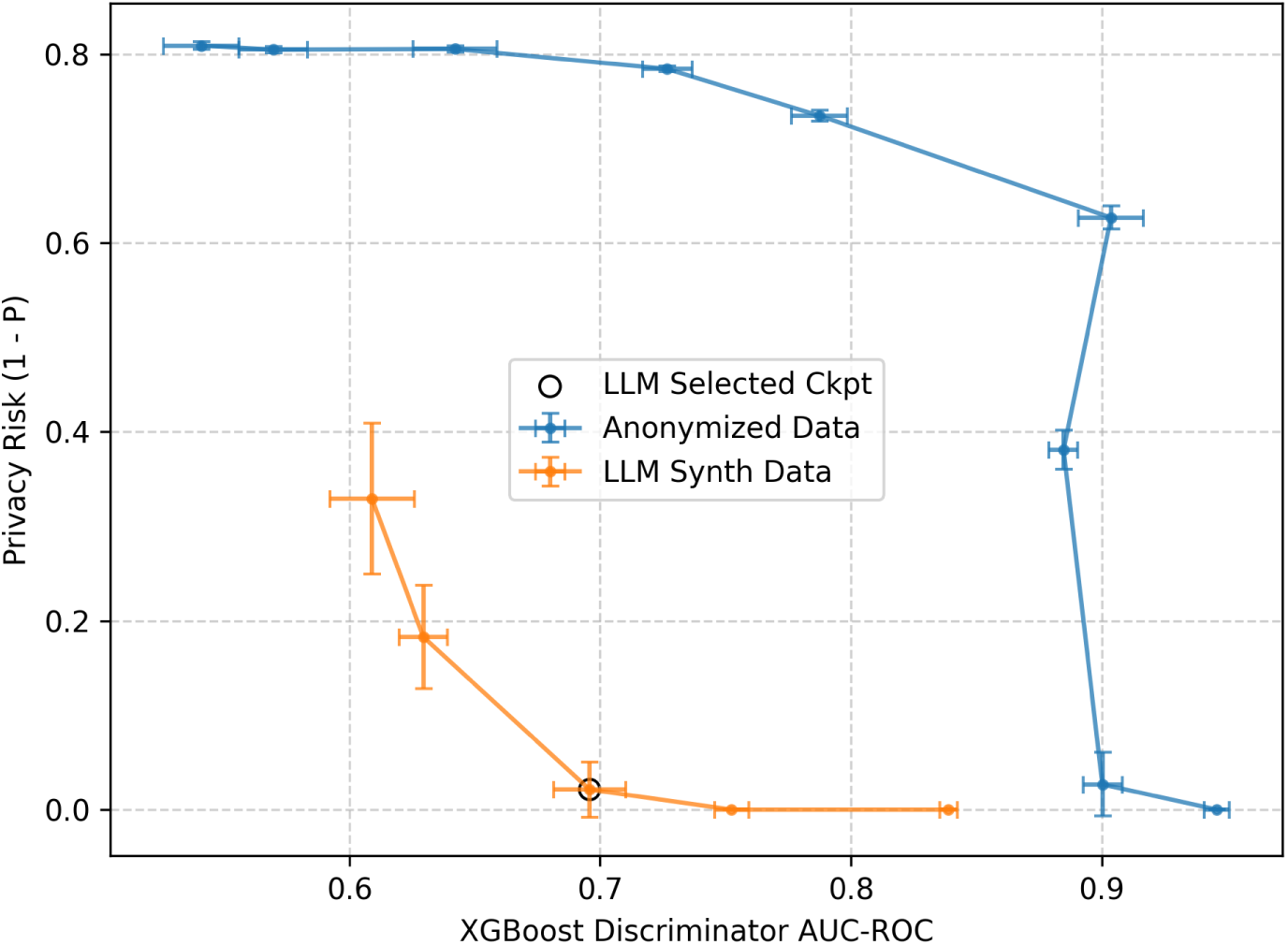
Utility-privacy trade-off for SAMS synthetic and anonymized datasets. The fidelity metric employed is the XGBoost discriminator AUC-ROC, where lower values indicate superior fidelity; Privacy risk, on the other hand was measured by (1-P) with k=1 and q=0.1. In this case as well, lower values are associated with a reduced risk. For the anonymized data (blue), increasing noise reduces both utility and privacy risk. For the synthetic data (orange), the circled point denotes the selected checkpoint, representing the best trade-off between data utility and privacy risk. Error bars represent 95% confidence intervals.

**Figure 2.**
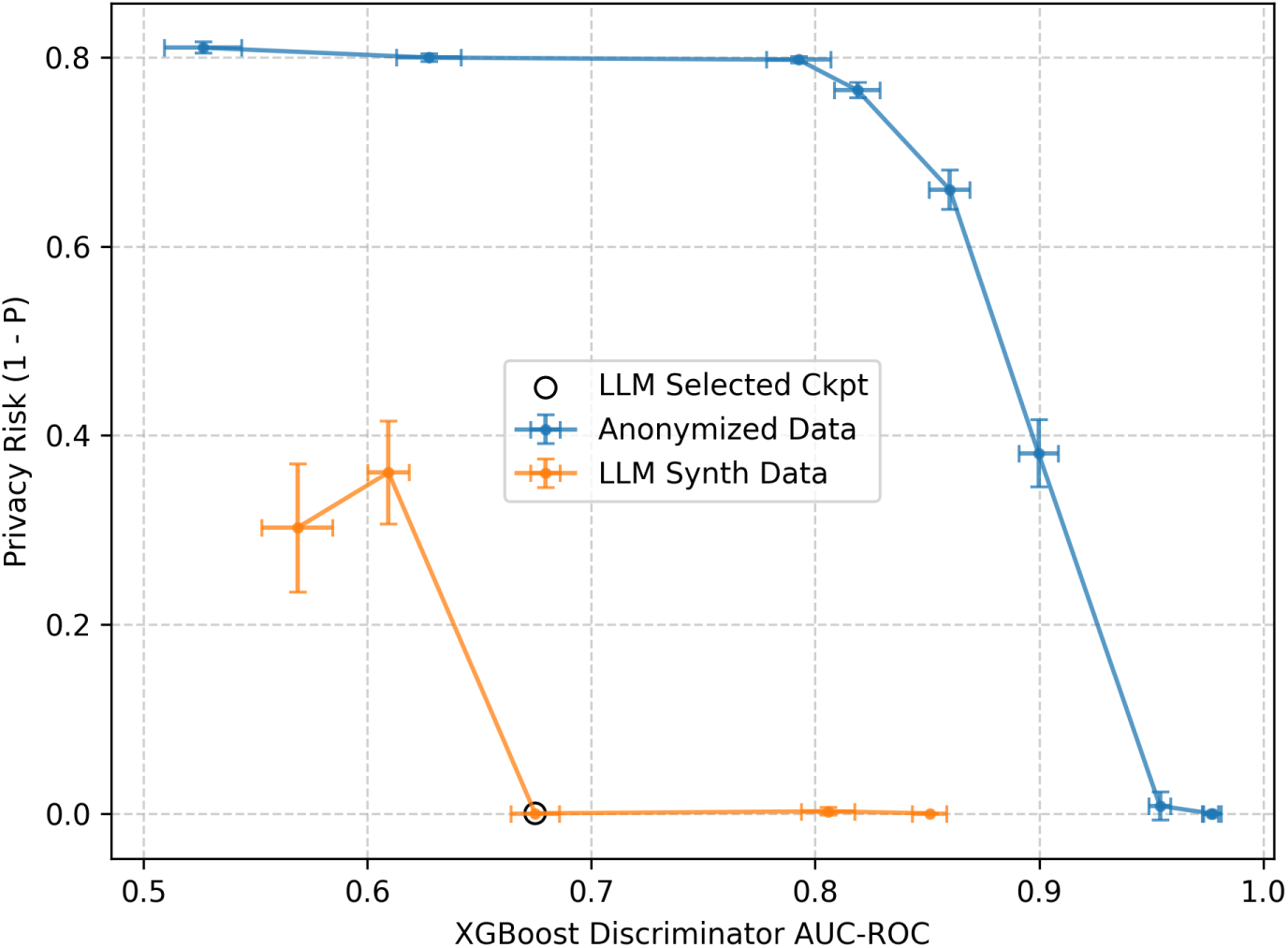
Utility-privacy trade-off for LDL-C synthetic and anonymized datasets. The fidelity metric employed is the XGBoost discriminator AUC-ROC, where lower values indicate superior fidelity; Privacy risk, on the other hand was measured by (1-P) with k=1 and q=0.1. In this case as well, lower values are associated with a reduced risk. For the anonymized data (blue), increasing noise reduces both utility and privacy risk. For the synthetic data (orange), the circled point denotes the selected checkpoint, representing the best trade-off between data utility and privacy risk. Error bars represent 95% confidence intervals.

**Figure 3.**
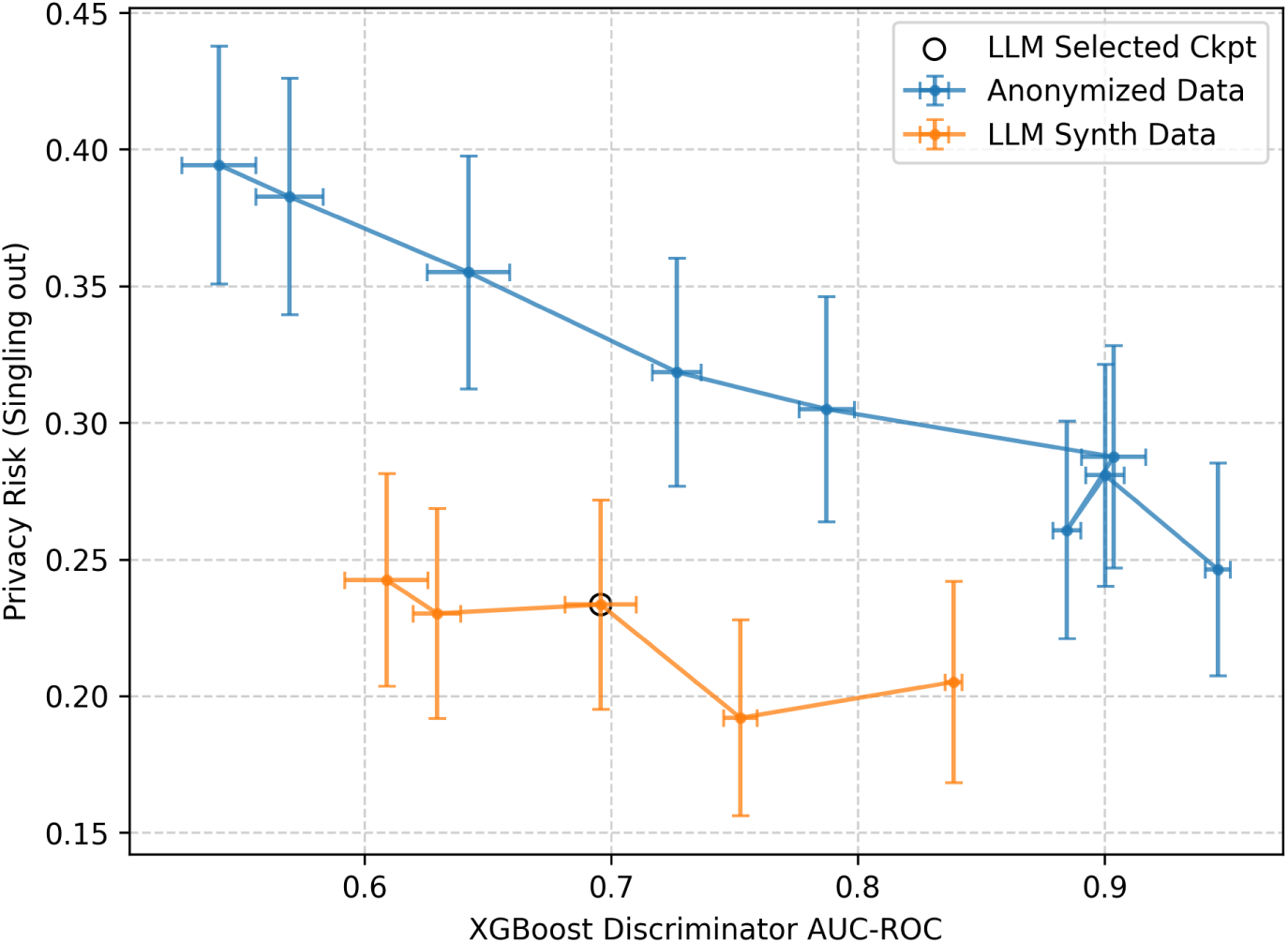
Utility-privacy trade-off for SAMS synthetic and anonymized datasets. The fidelity metric employed is the XGBoost discriminator AUC-ROC, where lower values indicate superior fidelity; Privacy risk, on the other hand was measured by the Anonymeter singling-out attack. For the anonymized data (blue), increasing noise reduces both utility and privacy risk. For the synthetic data (orange), the circled point denotes the selected checkpoint. Notably, the attack-based metric remains relatively stable across steps. Error bars represent 95% confidence intervals.

**Figure 4.**
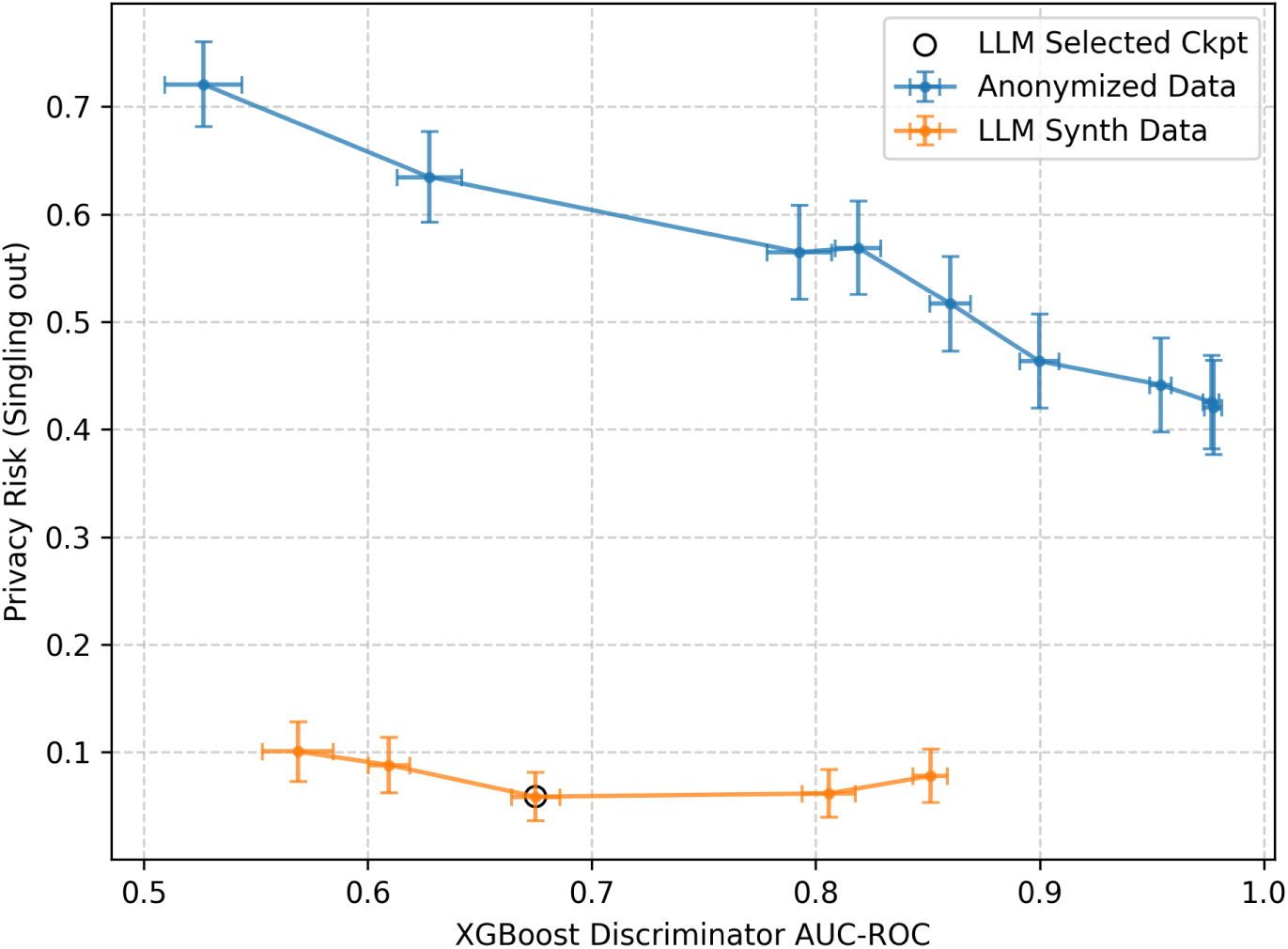
Utility-privacy trade-off for LDL-C synthetic and anonymized datasets. The fidelity metric employed is the XGBoost discriminator AUC-ROC, where lower values indicate superior fidelity; Privacy risk, on the other hand was measured by the Anonymeter singling-out attack. For the anonymized data (blue), increasing noise reduces both utility and privacy risk. For the synthetic data (orange), the circled point denotes the selected checkpoint. Notably, the attack-based metric remains relatively stable across steps. Error bars represent 95% confidence intervals.

**Figure 5.**
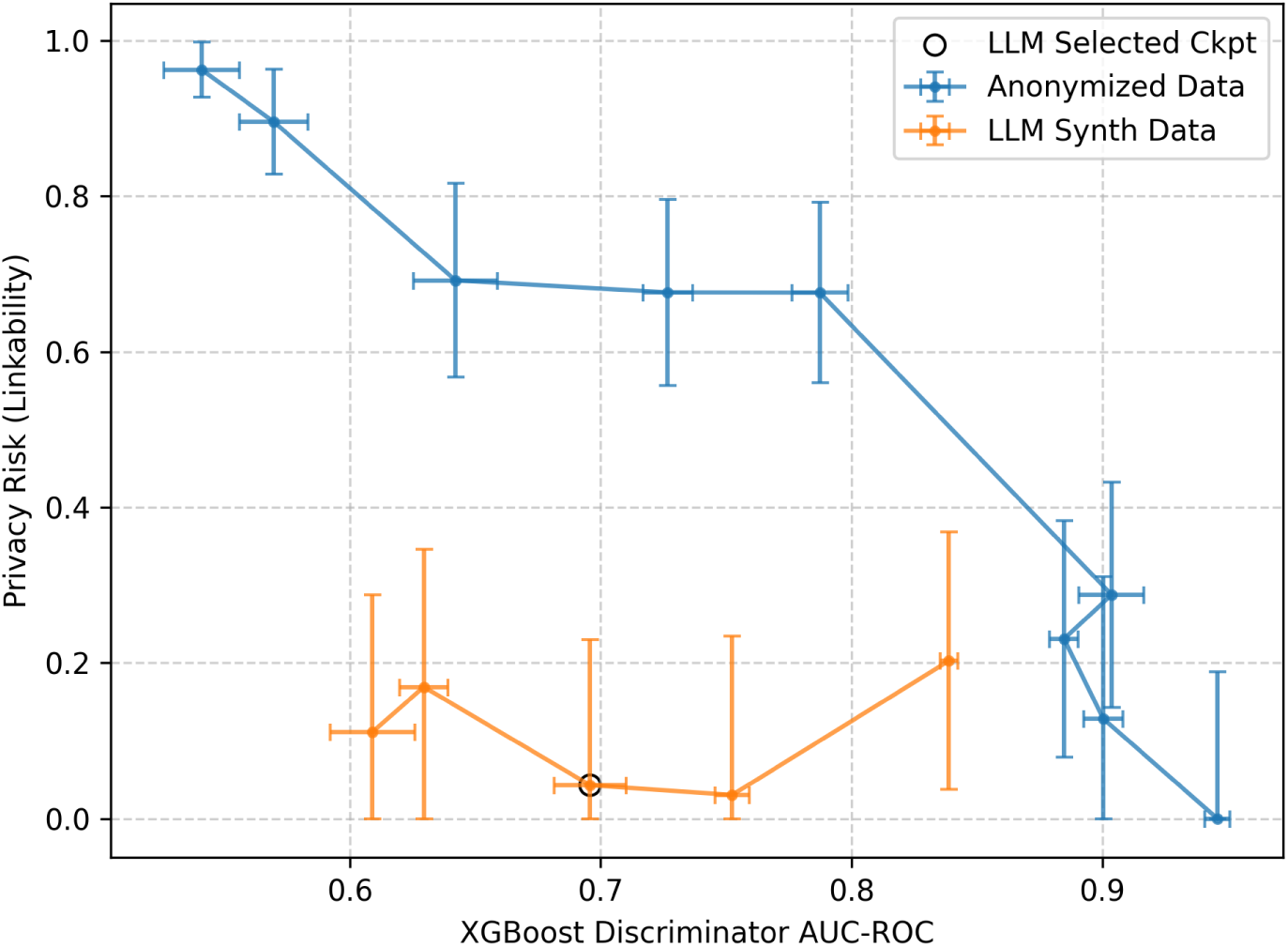
Utility-privacy trade-off for SAMS synthetic and anonymized datasets. The fidelity metric employed is the XGBoost discriminator AUC-ROC, where lower values indicate superior fidelity; Privacy risk, on the other hand was measured by the Anonymeter linkability attack. For the anonymized data (blue), increasing noise reduces both utility and privacy risk. For the synthetic data (orange), the circled point denotes the selected checkpoint. The attack-based metric remains relatively stable across steps. Error bars represent 95% confidence intervals.

**Figure 6.**
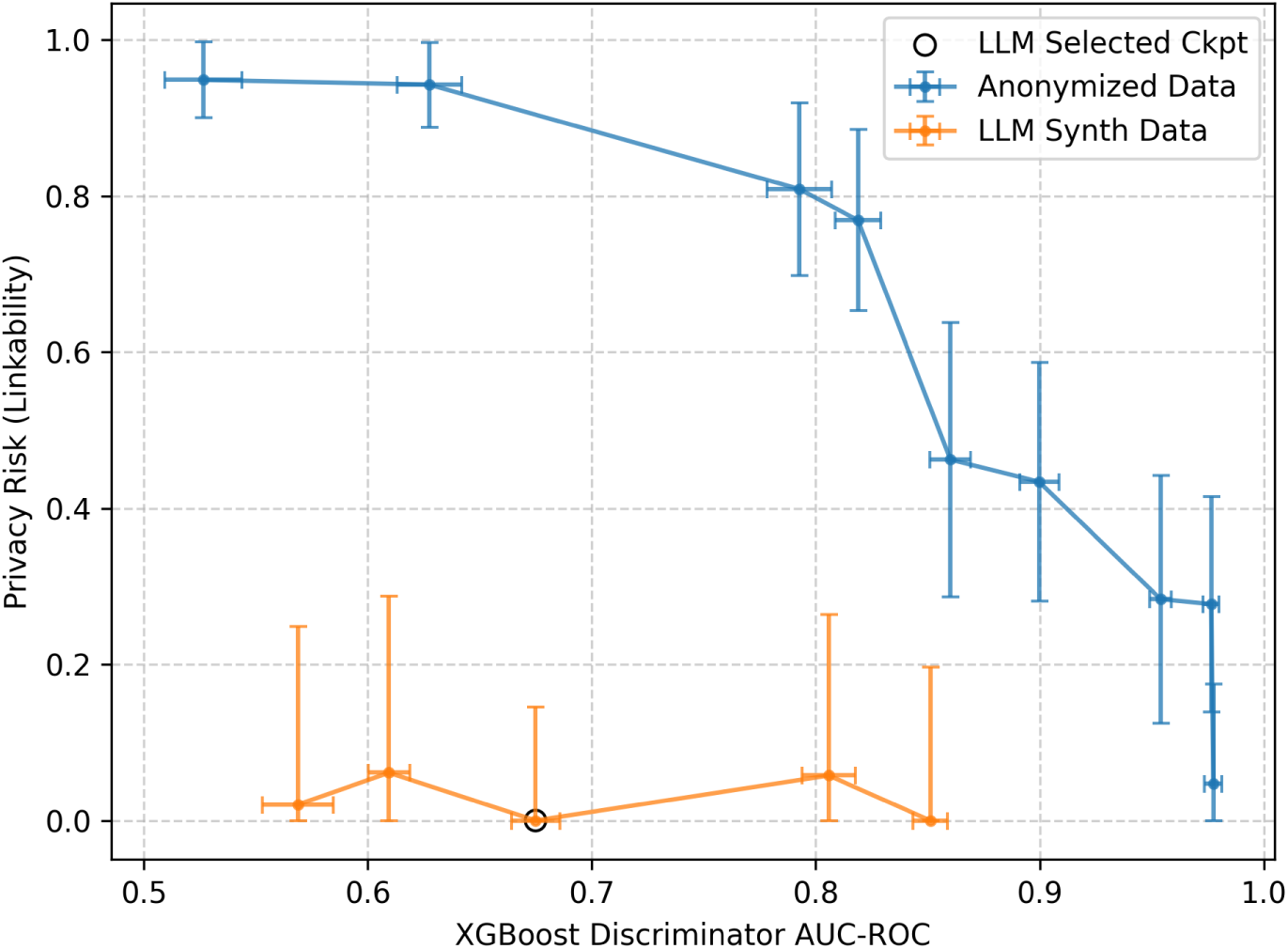
Utility-privacy trade-off for LDL-C synthetic and anonymized datasets. The fidelity metric employed is the XGBoost discriminator AUC-ROC, where lower values indicate superior fidelity; Privacy risk, on the other hand was measured by the Anonymeter linkability attack. For the anonymized data (blue), increasing noise reduces both utility and privacy risk. For the synthetic data (orange), the circled point denotes the selected checkpoint. The attack-based metric remains relatively stable across steps. Error bars represent 95% confidence intervals.

A detailed description of the parameters used in this section is provided in the Supplementary Materials.

To ensure the reliability of these metrics, we report error bars representing the 95% CI of the measured quantities:

- **XGBoost discriminator:** AUC-ROC is calculated across 10 iterations, utilizing a randomized 50% subsample of the data for each run.
- **Privacy Score:** Computed 10 times using different randomized partitions of the required subsets *R*_1_, *R*_2_, and *S*.
- **Anonymeter Attacks:** We utilize the confidence intervals natively generated by the tool.

Model checkpoints are saved every 100 training steps. For visual clarity, our plots display a representative subset of these checkpoints (at step 200, 600, 1000, 1400 and 1720). We observe a general trend where extended training enhances synthetic fidelity but concurrently increases the associated privacy risk.

The optimal configuration is identified at step 1000. While the attack-based metrics remain relatively stable across steps, the Privacy Score identifies this checkpoint as the final stage before a detectable degradation in privacy preservation occurs.

In all plots, we report the results for the anonymized baseline across varying noise parameters α (0.1, 0.2, 0.3, 0.4, 0.5, 0.6, 0.7, 0.8, and 0.9). At low noise levels (lower values of α), the anonymized data exhibits high fidelity but substantial privacy risk (occupying the upper-left quadrant). Conversely, as noise increases (higher values of α), both utility and risk diminish (moving toward the lower-right quadrant).

Across all measured dimensions, the synthetic data consistently outperforms the anonymized datasets, demonstrating a superior trade-off between privacy and clinical utility.

For completeness, Figure 7 and Figure 8 illustrate the Anonymeter inference attacks for two specific “secret” clinical columns. As for the linkability attacks, the limited size of the holdout set restricts the number of attacks to 100 for the SAMS dataset and 70 for the LDL-C dataset. This analysis was replicated across several “secret” columns, yielding consistent results regardless of the specific attribute selected. The inference risk for synthetic data does not present a strong trend throughout the training process, and exhibits high variance (noise). Consequently, this metric is excluded from the primary selection criteria. In contrast, for the anonymized baseline it follows the expected trend, where attack success rates decrease with injected noise. Comparing these results reinforces our conclusion that LLM-based synthetic data provides a more robust privacy-utility profile than classical perturbation methods.

**Figure 7.**
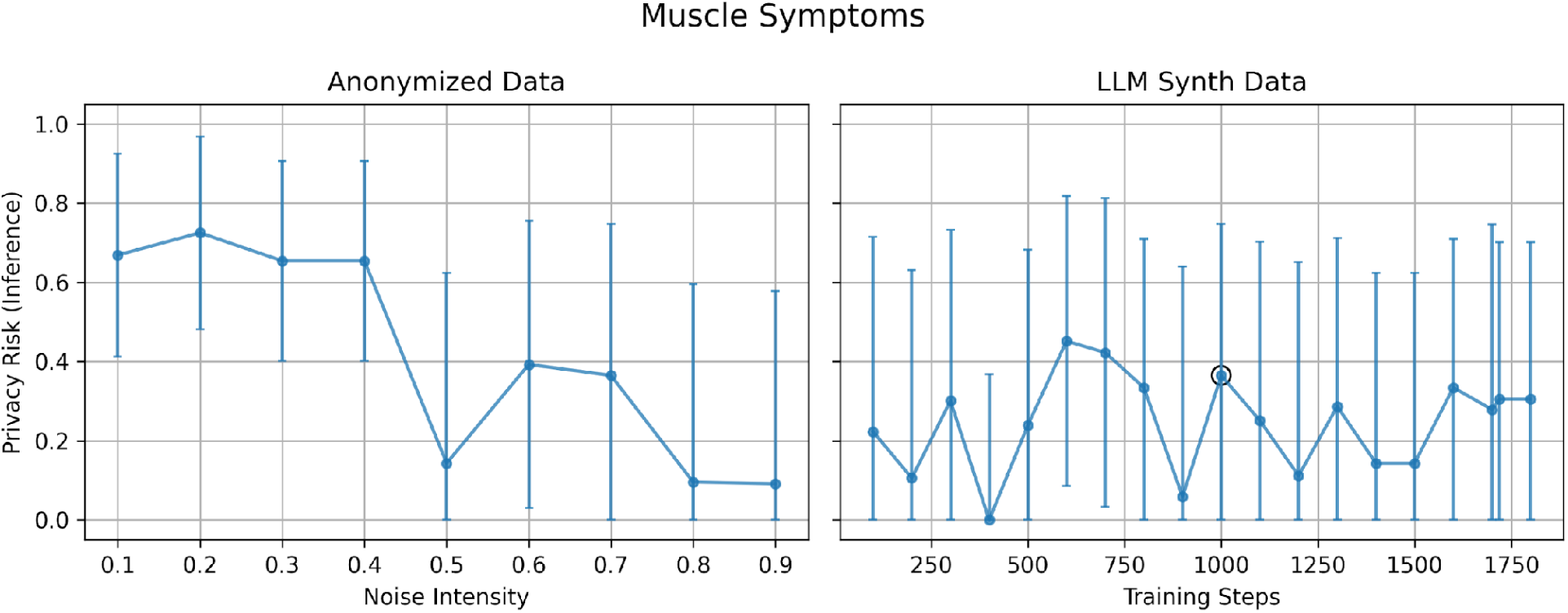
Privacy risk for SAMS synthetic and anonymized datasets. Privacy risk is measured by the Anonymeter inference attack, using “Statin Associated Muscle Symptoms” as the secret column. For anonymized data, results are reported as a function of the noise intensity α, while for synthetic data they are reported as a function of the training step. Error bars represent 95% confidence intervals.

**Figure 8.**
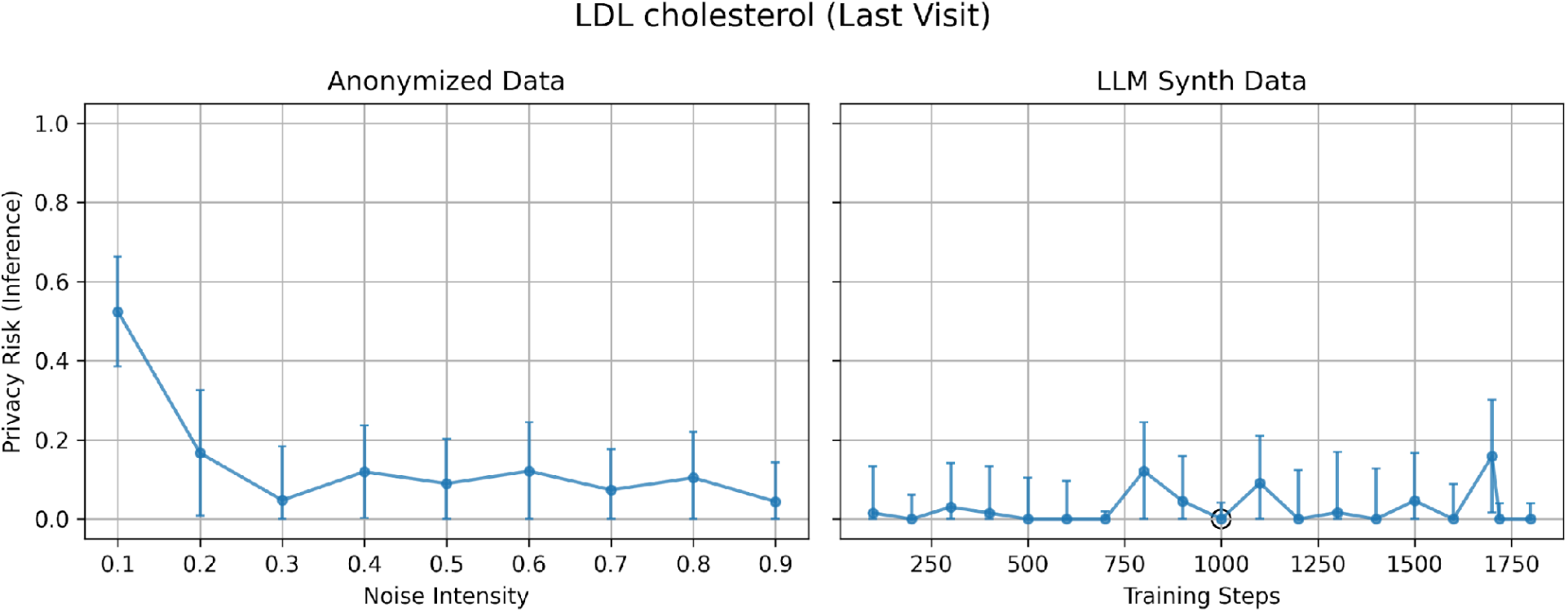
Privacy risk for LDL-C synthetic and anonymized datasets. Privacy risk is measured by the Anonymeter inference attack, using “LDL cholesterol (Last Visit)” as the secret column. For anonymized data, results are reported as a function of the noise intensity α, while for synthetic data they are reported as a function of the training step. Error bars represent 95% confidence intervals.

### Statistical analysis

#### SAMS use case

Among the 72 variables analyzed, only 7 (approximately 9.5%) showed a statistically significant difference in distribution between the real and synthetic datasets (Table 1). Continuous variables were assessed using the Wilcoxon rank-sum test, with p < 0.050 considered statistically significant, whereas categorical variables were evaluated using Jensen–Shannon Divergence (JSD), with JSD > 0.4 indicating a significant distributional difference. The distributions of the continuous variables in the real and synthetic datasets are presented in Figure 9.

**Figure 9.**
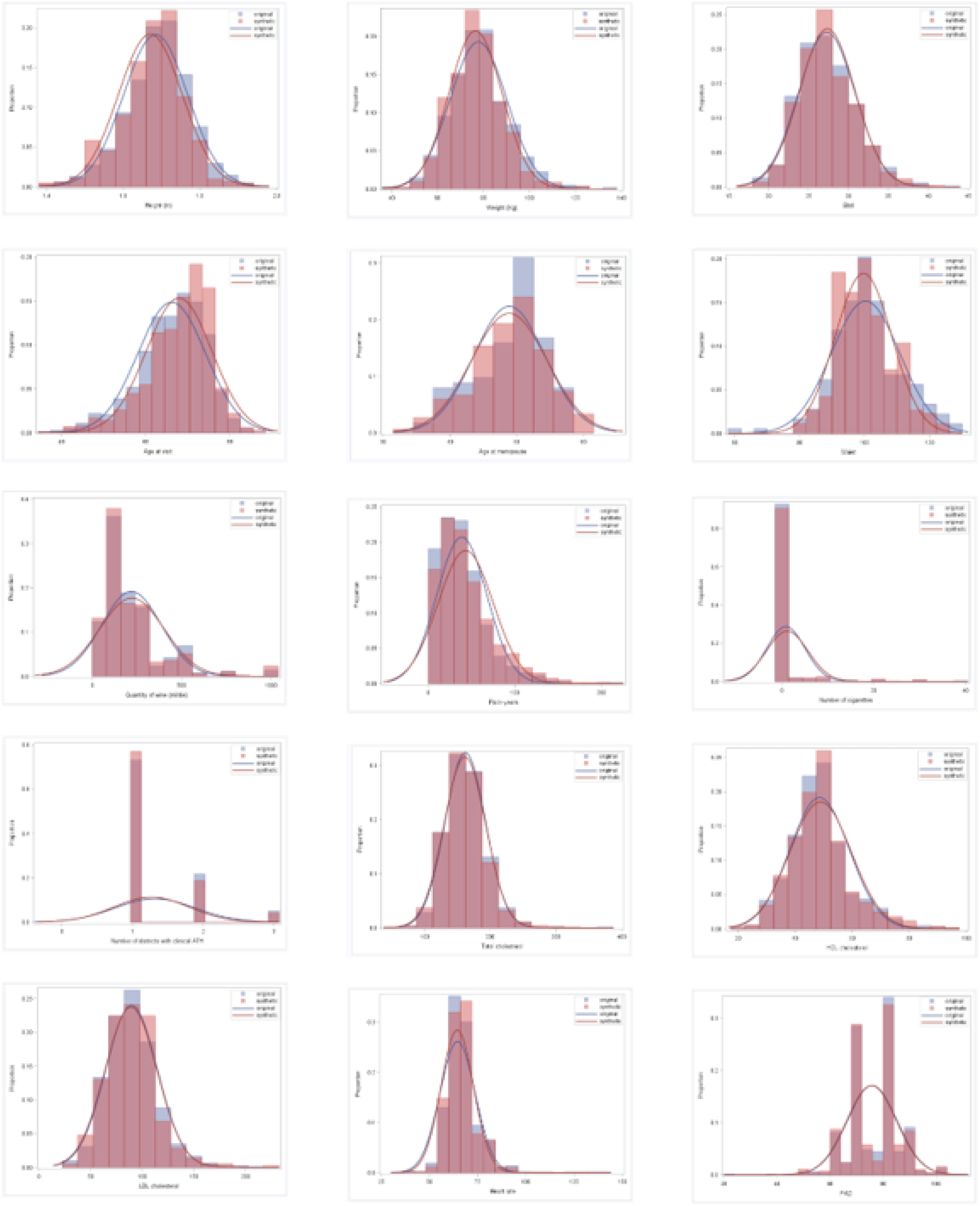
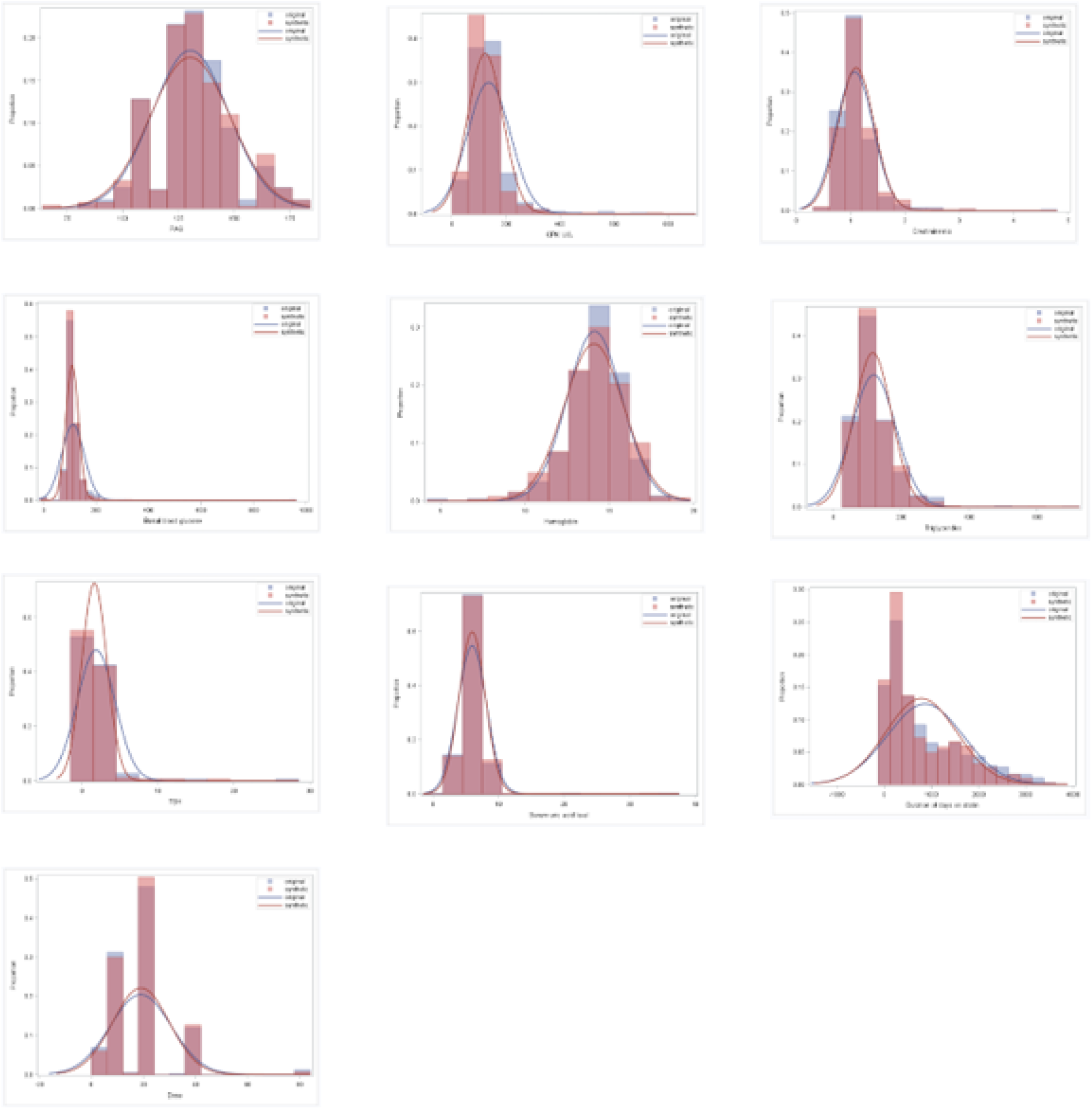
Distributions of continuous variables in real and synthetic SAMS datasets. Histograms and density curves are presented to compare the distribution of the 25 continuous variables in SAMS real (blue) and synthetic (red) datasets.

**Table 1.** Distribution of variables in the original and synthetic datasets for SAMS Use Case.

| Variables | Original dataset | Synthetic dataset | JSD<sup>h</sup> | p-value |
| --- | --- | --- | --- | --- |
| **Categorical** |  |  |  |  |
| Age ($\geq 65$) | 609 (60.90%) | 703 (70.30%) | 0.0071 | |
| Sex (Female) | 130 (13.00%) | 159 (15.90%) | 0.0012 |  |
| Education level |  |  | 0.0051 |  |
| Elementary school | 140 (14.49%) | 171 (17.87%) |  |  |
| Lower secondary school | 229 (23.71%) | 271 (28.32%) |  |  |
| Upper secondary school | 442 (45.76%) | 374 (39.08%) |  |  |
| University degree | 148 (15.32%) | 133 (13.90%) |  |  |
| Occupation |  |  | 0.0082 |  |
| Other | 150 (15.31%) | 148 (15.16%) |  |  |
| Housewife | 35 (3.57%) | 64 (6.56%) |  |  |
| Executive / Manager | 67 (6.84%) | 70 (7.17%) |  |  |
| Employee | 323 (32.96%) | 344 (35.25%) |  |  |
| Entrepreneur | 58 (5.92%) | 39 (4.00%) |  |  |
| Teacher | 30 (3.06%) | 33 (3.38%) |  |  |
| Self-employed | 121 (12.35%) | 134 (13.73%) |  |  |
| Freelance professional | 79 (8.06%) | 64 (6.56%) |  |  |
| Worker / Laborer | 117 (11.94%) | 80 (8.20%) |  |  |
| Marital status |  |  | 0.0042 |  |
| Married | 772 (80.84%) | 733 (78.23%) |  |  |
| Unmarried | 62 (6.49%) | 94 (10.03%) |  |  |
| Separated | 50 (5.24%) | 42 (4.48%) |  |  |
| Widowed | 63 (6.60%) | 58 (6.19%) |  |  |
| Social status |  |  | 0.0071 |  |
| Unemployed | 12 (1.20%) | 6 (0.60%) |  |  |
| Disabled | 8 (0.80%) | 7 (0.70%) |  |  |
| Employed | 387 (38.82%) | 307 (30.73%) |  |  |
| Retired | 576 (57.77%) | 668 (66.87%) |  |  |
| Physical activity outside of work |  |  | 0.0016 |  |
| Intense | 52 (5.22%) | 53 (5.34%) |  |  |
| Moderate | 578 (58.03%) | 566 (57.06%) |  |  |
| Low | 364 (36.55%) | 373 (37.60%) |  |  |
| Occupational physical activity |  |  | 0.0034 |  |
| Intense | 16 (1.61%) | 12 (1.21%) |  |  |
| Moderate | 152 (15.32%) | 109 (11.02%) |  |  |
| Low | 822 (82.86%) | 866 (87.56%) |  |  |
| Physical activity |  |  | 0.0010 |  |
| Low | 301 (30.53%) | 334 (33.84%) |  |  |
| Moderate | 621 (62.98%) | 588 (59.57%) |  |  |
| Intense | 64 (6.49%) | 65 (6.59%) |  |  |
| Smoking |  |  | 0.0102 |  |
| Former smoker | 623 (62.30%) | 705 (70.50%) |  |  |
| Current smoker | 74 (7.40%) | 94 (9.40%) |  |  |
| Never smoked | 303 (30.30%) | 201 (20.10%) |  |  |
| Spirits / Hard liquor | 66 (6.65%) | 40 (4.04%) | 0.0024 |  |
| BMI |  |  | 0.0002 |  |
| < 25 | 221 (27.15%) | 213 (25.60%) |  |  |
| 25–30 | 414 (50.86%) | 436 (52.40%) |  |  |
| ≥ 30 | 179 (21.99%) | 183 (22.00%) |  |  |
| Allergies | 189 (18.98%) | 158 (15.88%) | 0.0012 |  |
| Anxiety-depressive disorder | 124 (12.45%) | 105 (10.54%) | 0.0006 |  |
| History of cerebrovascular disease | 184 (18.42%) | 155 (15.53%) | 0.0011 |  |
| History of coronary disease | 945 (94.50%) | 929 (92.90%) | 0.0008 |  |
| Rheumatoid arthritis | 10 (1.01%) | 4 (0.40%) | 0.0010 |  |
| Asthma / COPD<sup>b</sup> / Emphysema | 133 (13.35%) | 109 (10.95%) | 0.0010 |  |
| Autoimmune Rheumatic Diseases | 50 (5.03%) | 47 (4.73%) | 0.0000 |  |
| Blood disorders | 61 (6.12%) | 61 (6.12%) | 0.0000 |  |
| Collagen diseases | 26 (2.61%) | 18 (1.81%) | 0.0005 |  |
| Chronic kidney disease | 37 (3.72%) | 16 (1.61%) | 0.0032 |  |
| Chronic liver disease | 120 (12.06%) | 114 (11.45%) | 0.0001 |  |
| Diabetes | 239 (23.97%) | 218 (21.89%) | 0.0004 |  |
| Type 2 diabetes | 239 (23.90%) | 218 (21.80%) | 0.0067 |  |
| Other forms of hyperglycemia | 126 (12.60%) | 74 (7.40%) |  |  |
| No diabetes | 635 (63.50%) | 708 (70.80%) |  |  |
| Dyslipidemia | 885 (88.59%) | 839 (83.98%) | 0.0032 |  |
| Other dyslipidemias | 70 (7.00%) | 106 (10.60%) | 0.0029 |  |
| Gallstones | 120 (12.06%) | 121 (12.15%) | 0.0000 |  |
| Hiatal hernia | 108 (10.85%) | 95 (9.54%) | 0.0003 |  |
| Hypercholesterolemia | 789 (78.90%) | 704 (70.40%) | 0.0069 |  |
| Hyperglycemia | 362 (36.35%) | 292 (29.32%) | 0.0040 |  |
| Hypertension | 705 (70.71%) | 712 (71.49%) | 0.0001 |  |
| Hypertriglyceridemia | 269 (26.90%) | 238 (23.80%) | 0.0009 |  |
| Hyperuricemia / Gout | 117 (11.76%) | 75 (7.53%) | 0.0037 |  |
| Infectious diseases | 204 (20.50%) | 199 (19.96%) | 0.0000 |  |
| Kidney stones | 214 (21.51%) | 194 (19.48%) | 0.0005 |  |
| Menopause (yes) | 116 (95.87%) | 150 (94.34%) | 0.0023 |  |
| Nephrotic syndrome | 2 (0.20%) | 1 (0.10%) | 0.0001 |  |
| Neoplasms / Cancer | 165 (16.57%) | 192 (19.28%) | 0.0009 |  |
| Overweight / Obesity | 383 (38.45%) | 323 (32.43%) | 0.0029 |  |
| Acute pancreatitis | 7 (0.70%) | 5 (0.50%) | 0.0001 |  |
| Thyroid disorders | 96 (9.65%) | 113 (11.35%) | 0.0006 |  |
| Thrombophilia | 18 (1.81%) | 4 (0.40%) | 0.0035 |  |
| Vascular disease in other districts | 190 (19.06%) | 187 (18.74%) | 0.0000 |  |
| Therapy modification (yes) | 154 (15.40%) | 187 (18.70%) | 0.0014 |  |
| Active ingredient |  |  | 0.0015 |  |
| Atorvastatin (Calcium Salt) | 299 (29.90%) | 312 (31.20%) |  |  |
| Fluvastatin (Sodium Salt) | 10 (1.00%) | 5 (0.50%) |  |  |
| Lovastatin | 2 (0.20%) | 2 (0.20%) |  |  |
| Pravastatin (Sodium Salt) | 47 (4.70%) | 54 (5.40%) |  |  |
| Rosuvastatin | 299 (29.90%) | 270 (27.00%) |  |  |
| Simvastatin | 343 (34.30%) | 357 (35.70%) |  |  |
| Statin intensity |  |  | 0.0004 |  |
| Low | 117 (11.78%) | 131 (13.25%) |  |  |
| Medium | 748 (75.33%) | 728 (73.61%) |  |  |
| High | 128 (12.89%) | 130 (13.14%) |  |  |
| **Numerical** |  |  |  |  |
| Height (m) | 1.68±0.08 | 1.67±0.08 |  | 0.001 |
| Weight (kg) | 77.78±12.41 | 76.54±11.53 |  | 0.038 |
| BMI<sup>a</sup> | 27.3±3.55 | 27.37±3.48 |  | 0.664 |
| Age at visit | 66.22±8.05 | 67.96±7.8 |  | <.0001 |
| Age at menopause | 48.88±5.36 | 48.88±5.67 |  | 0.854 |
| Waist circumference (periumbilical) | 100.3±10.53 | 99.68±8.7 |  | 0.360 |
| Wine consumption (ml per day) | 219.76±166.7<br>2 | 221.94±180.46 |  | 0.812 |
| Pack years | 38.84±28.85 | 43.24±31.78 |  | 0.017 |
| Number of cigarettes at visit | 0.87±4.18 | 1.15±4.55 |  | 0.096 |
| Number of districts with clinical atherosclerosis | 1.32±0.57 | 1.27±0.53 |  | 0.043 |
| Total cholesterol | 162.1±30.82 | 161.44±31.61 |  | 0.674 |
| HDL<sup>f</sup> cholesterol | 48.62±10.42 | 48.93±10.77 |  | 0.483 |
| Calculated LDL<sup>i</sup> cholesterol | 89.64±24.92 | 89.11±25.16 |  | 0.692 |
| HR<sup>g</sup> | 64.56±9.13 | 64.41±8.41 |  | 0.922 |
| DBP<sup>d</sup> | 75.87±9.34 | 75.68±9.34 |  | 0.731 |
| SBP<sup>j</sup> | 130.38±17.21 | 130.28±18 |  | 0.887 |
| CPK<sup>c</sup> (UI/L) | 136.18±79.8 | 122.66±65.39 |  | <.0001 |
| Serum creatinine | 1.07±0.34 | 1.1±0.33 |  | 0.113 |
| Fasting blood glucose | 111.78±43.02 | 108.94±24.14 |  | 0.573 |
| Hb<sup>e</sup> | 14.13±1.63 | 14.1±1.77 |  | 0.789 |
| Triglycerides | 119.07±64.65 | 116.09±55.39 |  | 0.934 |
| TSH<sup>k</sup> | 1.89±2.49 | 1.67±1.65 |  | 0.655 |
| Serum uric acid | 5.96±2.19 | 5.96±2 |  | 0.982 |
| Duration on statin (days) | 863.66±805.5<br>5 | 784.28±753.84 |  | 0.035 |
| Dose | 18.95±11.75 | 19.04±10.83 |  | 0.347 |

<sup>a</sup> Body Mass Index
<sup>b</sup> Chronic Obstructive Pulmonary Disease
<sup>c</sup> Creatine Phosphokinase
<sup>d</sup> Diastolic Blood Pressure
<sup>e</sup> Hemoglobin
<sup>f</sup> High Density Lipoprotein
<sup>g</sup> Heart Rate
<sup>h</sup> Jensen–Shannon Divergence
<sup>i</sup> Low Density Lipoprotein
<sup>j</sup> Systolic blood pressure
<sup>k</sup> Thyroid-stimulating hormone

To evaluate predictive consistency, a logistic regression model with stepwise selection and cross-validation was independently developed on both the real and synthetic datasets. Notably, two predictors—Therapy modification and Creatine phosphokinase—were consistently identified as significant across both models. In the real dataset, therapy modification showed an association with SAMS (OR = 0.10; 95% CI: 0.05–0.12), and a comparable effect was observed in the synthetic dataset (OR = 0.10; 95% CI: 0.06–0.16), with an 80% overlap in confidence intervals. Similarly, creatine phosphokinase showed a stable odds ratio of 1.01 in both datasets with a 73% overlap in confidence intervals (real: 1.01–1.01; synthetic: 1.01–1.01).

#### LDL-C use case

None of the analyzed variables exhibited statistically significant differences in distribution between the synthetic and real datasets (Table 2). Figure 10 shows the overlap of distributions of continuous variables between actual data and synthetic data.

**Figure 10.**
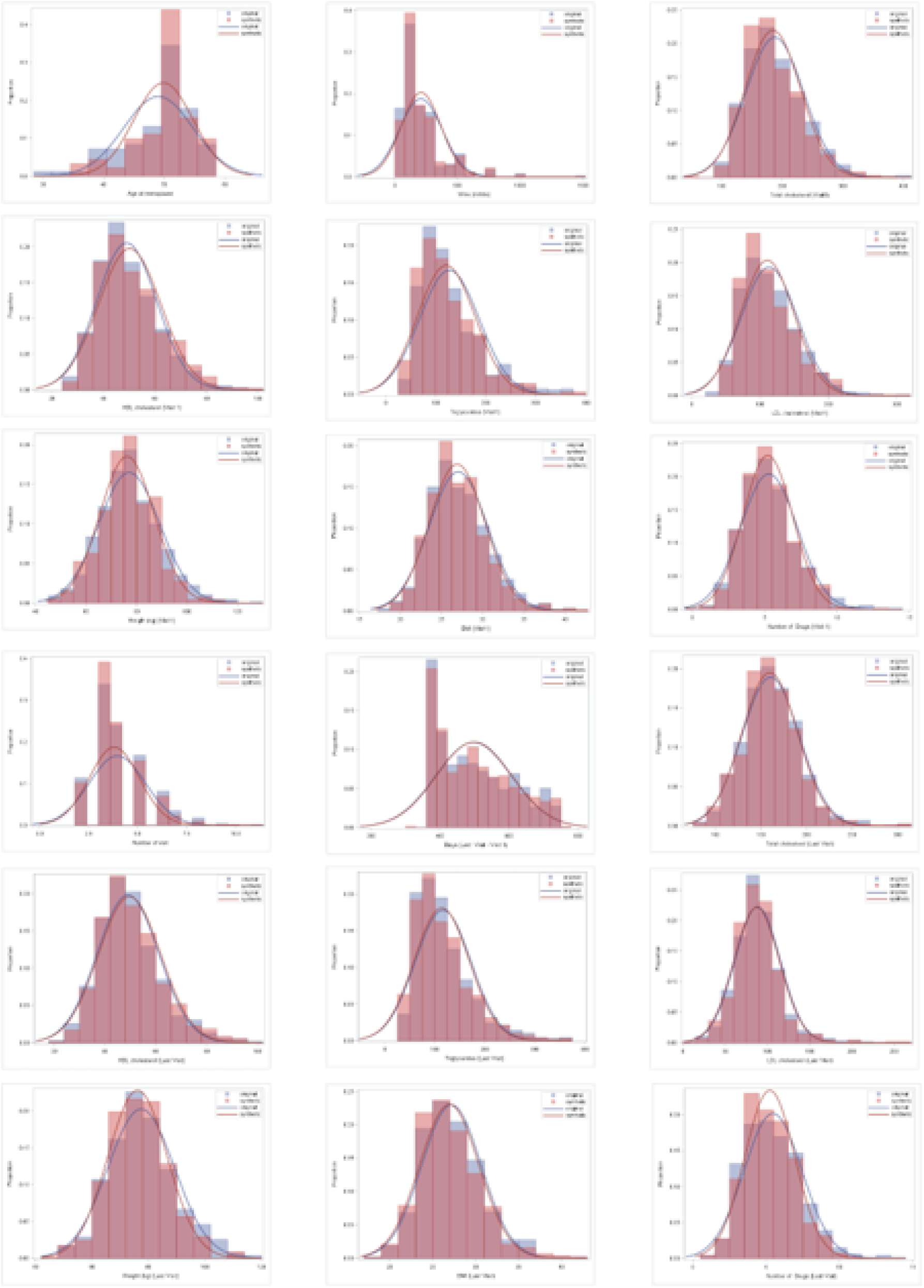
Distributions of continuous variables in LDL-C real and synthetic datasets. Histograms and density curves are presented to compare the distribution of the 18 continuous variables in LDL-C real (blue) and synthetic (red) datasets. None of the variables show a statistically significant difference in distribution across the two datasets.

**Table 2.** Distribution of variables in the original and synthetic datasets for LDL-C Use Case.

| Variables | Original dataset | Synthetic dataset | JSD<sup>d</sup> | p-value |
| --- | --- | --- | --- | --- |
| **Categorical** |  |  |  |  |
| Sex = Male | 604(86.8%) | 593(85.2%) | 0.0004 |  |
| Education Level |  |  | 0.0105 |  |
| Other | 3(0.4%) | 6(0.9%) |  |  |
| Primary School | 82(12.1%) | 63(9.4%) |  |  |
| Lower Secondary School | 155(23%) | 220(32.8%) |  |  |
| Upper Secondary School | 335(49.6%) | 280(41.8%) |  |  |
| University | 100(14.8%) | 101(15.1%) |  |  |
| Occupation |  |  | 0.0067 |  |
| Other | 100(14.6%) | 119(17.2%) |  |  |
| Housewife | 28(4.1%) | 31(4.5%) |  |  |
| Executive | 61(8.9%) | 50(7.2%) |  |  |
| Office Worker | 228(33.3%) | 255(37%) |  |  |
| Entrepreneur | 42(6.1%) | 29(4.2%) |  |  |
| Teacher | 20(2.9%) | 15(2.2%) |  |  |
| Self-employed | 73(10.7%) | 85(12.3%) |  |  |
| Freelancer | 68(9.9%) | 54(7.8%) |  |  |
| Manual Worker | 65(9.5%) | 52(7.5%) |  |  |
| Marital Status |  |  | 0.0023 |  |
| Other | 7(1%) | 6(0.9%) |  |  |
| Married | 549(81.8%) | 537(79.9%) |  |  |
| Not Married | 41(6.1%) | 54(8%) |  |  |
| Separated | 36(5.4%) | 46(6.8%) |  |  |
| Widowed | 38(5.7%) | 29(4.3%) |  |  |
| Occupational Physical Activity |  |  | 0.0014 |  |
| High | 17(2.5%) | 12(1.7%) |  |  |
| Moderate | 91(13.2%) | 78(11.3%) |  |  |
| Low | 583(84.4%) | 603(87%) |  |  |
| Leisure-time Physical Activity |  |  | 0.0008 |  |
| High | 35(5%) | 29(4.2%) |  |  |
| Moderate | 422(60.8%) | 420(60.7%) |  |  |
| Low | 236(34%) | 241(34.8%) |  |  |
| Smoking Status |  |  | 0.0044 |  |
| Former Smoker | 443(63.8%) | 493(71%) |  |  |
| Current Smoker | 32(4.6%) | 23(3.3%) |  |  |
| Never Smoker | 219(31.6%) | 178(25.6%) |  |  |
| Rheumatoid Arthritis | 4(0.6%) | 2(0.3%) | 0.0011 |  |
| Autoimmune Rheumatic Diseases | 34 (4.9%) | 24 (3.5%) | 0.0017 |  |
| Blood Disorders = Yes | 31(4.5%) | 21(3%) | 0.0010 |  |
| Thrombophilia | 14(2%) | 11(1.6%) | 0.0009 |  |
| Asthma / COPD<sup>b</sup> / Emphysema | 82(11.8%) | 64(9.2%) | 0.0020 |  |
| Allergies | 156(22.4%) | 158(22.7%) | 0.0007 |  |
| Thyroid Disorders | 69(9.9%) | 54(7.8%) | 0.0018 |  |
| Other Endocrine Disorders | 8(1.2%) | 4(0.6%) | 0.0014 |  |
| Infectious Diseases | 151(21.7%) | 147(21.1%) | 0.0008 |  |
| Cancers | 110(15.8%) | 127(18.2%) | 0.0015 |  |
| Peptic Ulcer Disease | 69(9.9%) | 65(9.3%) | 0.0008 |  |
| Hiatal Hernia | 65(9.4%) | 61(8.8%) | 0.0008 |  |
| Chronic Liver Disease | 84(12.1%) | 99(14.2%) | 0.0014 |  |
| Acute Pancreatitis | 4(0.6%) | 2(0.3%) | 0.0011 |  |
| Gallstones (Cholelithiasis) | 80(11.5%) | 88(12.6%) | 0.0009 |  |
| Acute renal failure | 7(1%) | 4(0.6%) | 0.0012 |  |
| Chronic renal failure | 31(4.5%) | 28(4%) | 0.0008 |  |
| Nephrolithiasis | 154(22.2%) | 155(22.3%) | 0.0007 |  |
| Anxious-depressive syndrome | 87(12.5%) | 86(12.4%) | 0.0014 |  |
| Overweight/Obesity | 278(39.9%) | 260(37.4%) | 0.0005 |  |
| Hyperuricemia/Gout | 81(11.6%) | 47(6.8%) | 0.0052 |  |
| Hyperglycemia | 243(34.9%) | 238(34.2%) | 0.0000 |  |
| Hypertension | 471(67.7%) | 505(72.6%) | 0.0021 |  |
| Dyslipidemia | 598(86.3%) | 652(93.9%) | 0.0122 |  |
| History of coronary artery disease | 662(95.1%) | 668(96%) | 0.0003 |  |
| Coronary condition type |  |  | 0.0104 |  |
| Unstable angina | 51(7.8%) | 25(3.8%) |  |  |
| Unclassified angina | 57(8.7%) | 46(6.9%) |  |  |
| Stable angina | 216(33.1%) | 232(35%) |  |  |
| Negative diagnostic coronary angiography | 1 (0.2%) | 5 (0.8%) |  |  |
| Positive diagnostic coronary angiography | 58 (8.9%) | 55 (8.3%) |  |  |
| Manifest myocardial infarction | 165(25.3%) | 192(29%) |  |  |
| Unrecognized myocardial infarction | 35(5.4%) | 40(6%) |  |  |
| Silent ischemia | 63(9.6%) | 60(9.1%) |  |  |
| Surgical revascularization | 2(0.3%) | 5(0.8%) |  |  |
| Endoluminal revascularization | 5(0.8%) | 2(0.3%) |  |  |
| History of cerebrovascular disease | 118(17%) | 108(15.5%) | 0.0003 |  |
| Vascular disease in other districts | 115(16.5%) | 62(8.9%) | 0.0095 |  |
| Initial statin intensity level |  |  | 0.0060 |  |
| No statin | 201(28.9%) | 155(22.3%) |  |  |
| Low intensity | 45(6.5%) | 36(5.2%) |  |  |
| Moderate intensity | 387(55.6%) | 420(60.3%) |  |  |
| High intensity | 63(9.1%) | 85(12.2%) |  |  |
| Final statin intensity level |  |  | 0.0029 |  |
| No statin | 73(10.5%) | 66(9.5%) |  |  |
| Low intensity | 69(9.9%) | 56(8%) |  |  |
| Moderate intensity | 449(64.5%) | 439(63.1%) |  |  |
| High intensity | 105(15.1%) | 135(19.4%) |  |  |
| Change in statin intensity |  |  | 0.0028 |  |
| Decreased | 45(6.5%) | 65(9.3%) |  |  |
| Unchanged | 439(63.1%) | 445(63.9%) |  |  |
| Increased | 212(30.5%) | 186(26.7%) |  |  |
| **Numerical** |  |  |  |  |
| Age at menopause | 49.01±5.7 | 49.86±4.86 |  | 0.366 |
| Wine (ml/die) | 207.15±170.4 | 209.25±157.58 |  | 0.667 |
| Total cholesterol (First Visit) | 187.81±47.68 | 184.8±45.36 |  | 0.221 |
| HDL<sup>c</sup> cholesterol (First Visit) | 49.05±11.65 | 50.18±12.12 |  | 0.096 |
| Triglycerides (First Visit) | 127.25±59.84 | 121.16±57.57 |  | 0.052 |
| LDL<sup>e</sup> cholesterol (First Visit) | 113.31±41.29 | 110.67±39.25 |  | 0.172 |
| Weight (kg) (First Visit) | 77.12±12.11 | 76.28±10.79 |  | 0.397 |
| BMI<sup>a</sup> (First Visit) | 27.03±3.56 | 26.89±3.37 |  | 0.427 |
| Number of Drugs (First Visit) | 5.28±1.95 | 5.21±1.72 |  | 0.853 |
| Number of visits | 3.97±1.44 | 3.82±1.28 |  | 0.087 |
| Days (Last Visit - First Visit) | 497.25±109.68 | 498.84±109.25 |  | 0.743 |
| Total cholesterol (Last Visit) | 160.57±31.64 | 159.72±30.69 |  | 0.653 |
| HDL<sup>c</sup> cholesterol (Last Visit) | 49.08±12.09 | 49.72±12.22 |  | 0.359 |
| Triglycerides (Last Visit) | 116.59±55.88 | 113.84±55.02 |  | 0.306 |
| LDL<sup>e</sup> cholesterol (Last Visit) | 88.17±26.87 | 87.1±27.02 |  | 0.459 |
| Weight (kg) (Last Visit) | 77.47±11.8 | 76.2±10.48 |  | 0.145 |
| BMI<sup>a</sup> (Last Visit) | 27.19±3.47 | 26.94±3.46 |  | 0.150 |
| Number of Drugs (Last Visit) | 5.46±1.98 | 5.25±1.71 |  | 0.109 |

<sup>a</sup> Body Mass Index
<sup>b</sup> Chronic Obstructive Pulmonary Disease
<sup>c</sup> High Density Lipoprotein
<sup>d</sup> Jensen–Shannon Divergence
<sup>e</sup> Low Density Lipoprotein

A Generalized Linear Model (GLM) was employed to assess the association between LDL-C change, clinical factors, and therapeutic management. In both cases, pharmacological intensity was identified as the most significant predictor of LDL-C reduction (β = 16.4 (Standard Error = 1.3) for real data; β = 13.1 (Standard Error = 1.2) for Synthetic data).

A bootstrap analysis with 5,000 samples indicated a substantial overlap between the parameter distributions from the real and synthetic datasets, further confirming the robustness of the findings (Figure 11).

**Figure 11.**
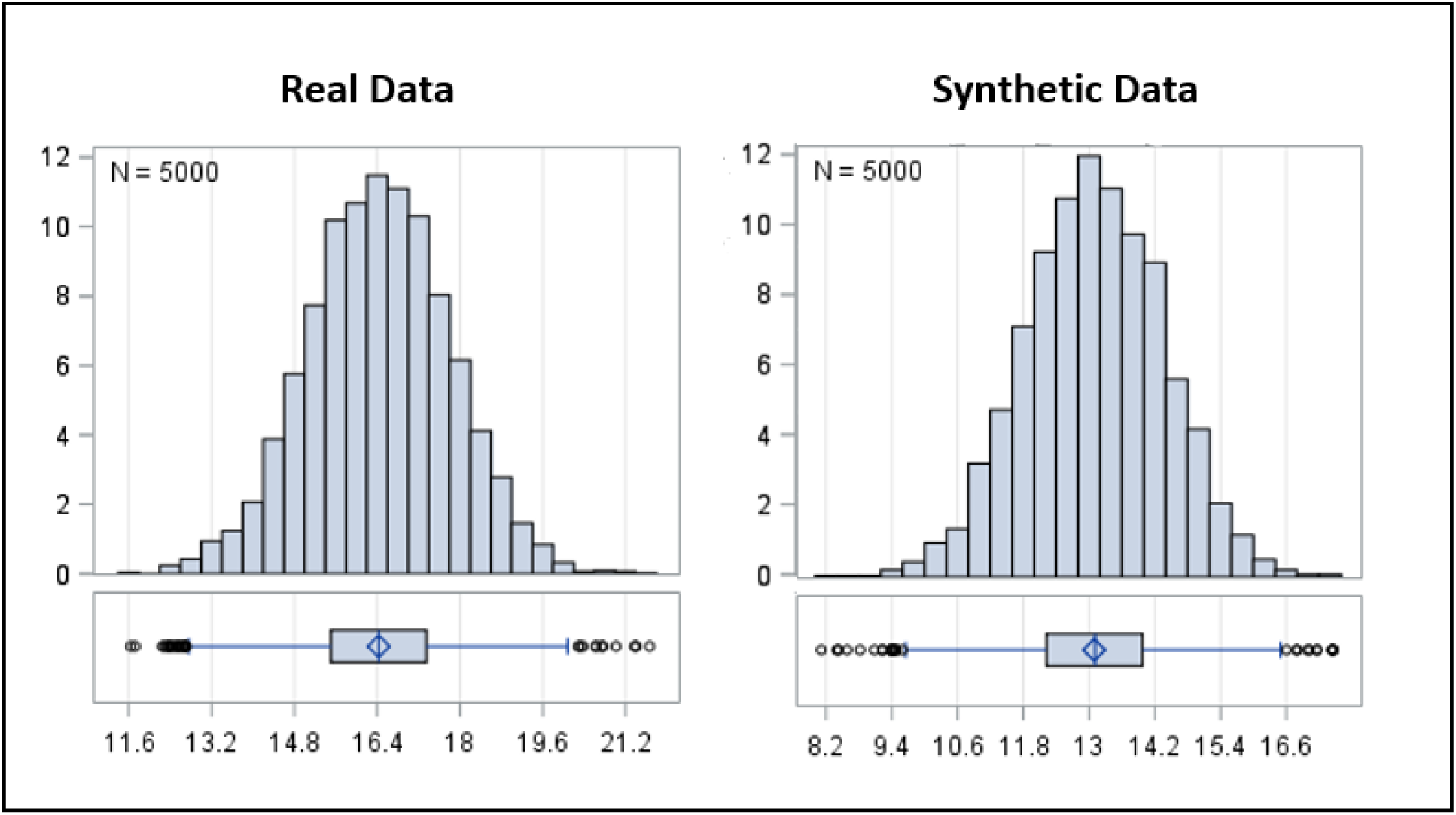
Real vs. Synthetic data: Bootstrap distribution of Beta. A generalized linear model (GLM) was used to evaluate the association between LDL-C levels and statin intensity, identified as the strongest predictor of LDL-C reduction. Parameter estimates were obtained from 5,000 bootstrap resamples of both the real and synthetic datasets. The figure shows a substantial overlap between the distributions of the estimated coefficients.

## Discussion

In this study, we evaluated the utility of AI-generated synthetic data within the context of secondary cardiovascular prevention for patients with dyslipidemia. To address the inherent challenges of limited clinical sample sizes, we implemented a generative framework based on Large Language Models (LLMs), utilizing constrained decoding to ensure strict adherence to the original tabular schema. The LLM was fine-tuned directly on the original clinical datasets within the Centro Cardiologico Monzino’s secure infrastructure, a strategy that allowed the model to learn the necessary statistical distributions without exposing sensitive patient records to external environments.

A key strength of this approach lies in its ability to harness transfer learning: by leveraging the general clinical priors acquired by the LLM during its pre-training phase, the model can effectively contextualize the specific statistical nuances of the target clinical datasets during fine-tuning. Furthermore, the architectural flexibility of the LLM permitted a multitask training strategy, wherein a single generative model was trained on both datasets simultaneously. This allowed the model to develop a more robust internal representation by learning from shared clinical features across both cohorts.

To identify the optimal generative model, we employed a multi-dimensional selection framework that rigorously evaluated the trade-off between statistical accuracy (fidelity) and privacy protection. We identified the optimal training checkpoint that maximized data fidelity while strictly adhering to predefined privacy thresholds. Furthermore, we benchmarked these results against a baseline of classically anonymized data. This comparison demonstrates that LLM-based synthetic data provides a significantly superior privacy-utility trade-off, offering a more robust solution for secure clinical data sharing than traditional perturbation methods.

To assess the practical utility of the generated data, the selected synthetic datasets were subjected to a downstream statistical analysis, the results of which were benchmarked against identical analyses performed on the original records. This comparative evaluation served to validate whether the synthetic cohorts could function as a privacy-preserving proxy for real-world data, without compromising the integrity of clinical conclusions.

Deviations in descriptive statistics were within an acceptable range, slightly higher than the theoretical expectation of random sampling variability.

A regression analysis was performed on the synthetic data and was compared with the same models applied on the real data. Using stepwise selection and cross validation, the identified predictors were found to match between the synthetic and real data. This suggests that synthetic data can serve as a reliable proxy for statistical inference, preserving core relationships among clinical variables.

This study has several limitations that should be considered when interpreting the findings. First, the retrospective study design is inherently subject to limitations related to the completeness of data originally collected for clinical rather than research purposes. Any data entry errors, missing information, or selection biases present in the original dataset may have been propagated to the generated synthetic data.

Second, the study was conducted at a single center, which may limit the representativeness of the study population compared with other clinical settings. Site-specific demographic, organizational, and healthcare delivery characteristics may influence both the distribution of variables and the performance of the synthetic data generation models. Consequently, the generalizability of the findings to other healthcare institutions or patient populations should be confirmed through multicenter validation studies.

Another limitation relates to the relatively modest size of the datasets used in the two use cases (700 and 1,000 patients, respectively). Although these sample sizes are adequate for evaluating the statistical fidelity and analytical utility of the synthetic data, they may not be sufficient to accurately capture underrepresented clinical subgroups.

Finally, the study did not assess the operational impact of using synthetic data in real-world settings, such as clinical research, algorithm development, or inter-institutional data sharing. Prospective and multicenter studies will be necessary to further validate the robustness of the findings and to determine the practical value of synthetic data across different application domains.

## Conclusions and outlook

In conclusion, our findings indicate that LLM-generated synthetic data can serve as an effective privacy-preserving proxy for real-world clinical datasets, maintaining the key statistical structures and inferential conclusions of the original cohorts while offering a superior privacy–utility trade-off compared with traditional anonymization approaches.

These findings support the broader use of synthetic data as a privacy-preserving alternative for exploratory analyses, hypothesis generation, and model development in clinical research. Nonetheless, researchers should remain mindful of the potential for conservative bias in effect estimates derived from synthetic data.

## Data Availability

The original data are not publicly available.
The synthetic data generated will be made available to the researchers upon request.

https://github.com/aindo-com/monzino-sams-ldl

## Abbreviations

AI: Artificial Intelligence
AIA: Attribute Inference Attacks
AUC: Area Under Curve
CI: Confidence Interval
DCR: Distance to Closest Record
GDPR: General Data Protection Regulation
GLM: General Linear Model
HIPAA: Health Insurance Portability and Accountability Act
JSD: Jensen–Shannon Divergence
LDL-C: Low-Density Lipoprotein Cholesterol
LLM: Large Language Model
MIA: Membership Inference Attacks
ML: Machine Learning
OR: Odds Ratio
ROC: Receiver Operating Characteristic
RWD: Real-World Data
SAMS: Statin-Associated Muscle Symptoms

