## Supplementary Materials for "Evaluation of AI-Generated Synthetic Data for Clinical Research in Secondary Cardiovascular Prevention among Dyslipidemia Patients"

### Table of contents

Table of contents 1

Supplemental Methods 3

Technical details on synthetic data generation 3

Data preprocessing 3

Target columns 4

SAMS dataset 4

LDL-C dataset 5

Generation 5

Structured generation 5

Enforcing of fuzzy bounds 6

Leveraging pre-trained semantic knowledge 7

Fine-tuning 8

Training datasets and data partitioning 8

Hyperparameters and configuration 9

Technical details on model selection 10

Details on the XGBoost discriminator 10

The computation of the *Privacy Score* 12

Anonymeter attacks 13

Classical anonymization 15

References 18

Supplemental tables 19

### Supplemental Methods

#### Technical details on synthetic data generation

##### Data preprocessing

To ensure that the synthetic data strictly adheres to the original clinical dataset's schema, we utilize constrained (structured) generation techniques. These methods guide the LLM's decoding process to follow a predefined format, preventing the model from producing unstructured or hallucinated text that would break the tabular structure. To facilitate this, the clinical records are serialized into a JSON (JavaScript Object Notation) structure, where each dataset column was mapped to a specific JSON property.

The source datasets comprise four primary data types:

- **Categorical:** Multi-class variables, including binary (1/0) indicators, representing the presence or absence of specific conditions.
- **Integer:** Discrete numerical values and counts.
- **Numerical:** Continuous decimal values.
- **Datetime:** Temporal values standardized to a uniform ISO format during preprocessing.

Because these types are natively supported by JSON, no extensive feature engineering is required beyond the standardization of datetime formats. To represent missing values (NA) in the data, the native JSON null constant was used.

##### Target columns

Both datasets include primary target variables intended for downstream statistical analysis. In the SAMS dataset, the target is a binary indicator representing the presence or absence of muscle symptoms. In the LDL-C dataset, the target is defined as the relative change in LDL-C and it is derived from two distinct columns in the original dataset—the initial clinical visit value and the final visit value.

While the primary objective of high-quality synthetic data is the faithful replication of the original data's statistical distribution, it is standard practice to implement specific methodological refinements to ensure the preservation of critical clinical correlations.

##### SAMS dataset

For the SAMS dataset, given the inherent class imbalance of the target columns, stochastic fluctuations during generation could potentially bias downstream statistical analyses—a possibility exacerbated by the relatively small sample size of the cohort. To mitigate this, we employ conditional generation to preserve the marginal frequencies of the target variables.

In this framework, the target value is treated as a "context" and the remaining attributes as a "completion". During inference, the JSON-serialized context is provided as a prompt, and constrained decoding enforces the generation of the completion schema. This strategy does not compromise privacy, as the target variables are neither identifiers nor quasi-identifiers, and their aggregate distribution is not considered sensitive information. To support this capability, the fine-tuning objective must be specifically adapted: each training record is split into context and completion segments. Only the completion segment is used to calculate the training loss, ensuring the model learns to generate the desired attributes, conditioned on the provided context.

##### LDL-C dataset

For the LDL-C dataset, we replace the final LDL-C value with the calculated LDL-C change (the delta between the first and last visits) prior to training. By including this derived feature directly in the training set, we encourage the model to capture the complex correlations between treatment response and other baseline variables. Following the generation process, the synthetic patient's final LDL-C value is deterministically recomputed by adding the generated delta to the LDL-C value at the initial visit.

#### Generation

##### Structured generation

As previously noted, we employ structured generation to ensure that the synthetic output strictly adheres to the original clinical schema. Our generation pipeline utilizes vLLM[1] for high-throughput model serving, with guidance[2] as the structured output backend.

This approach requires the definition of a comprehensive JSON schema that dictates the properties of each generated record. Each dataset column is mapped to a JSON property with specific type constraints and validation metadata. The mapping logic is summarized in Supplemental Table 1 and detailed below:

- **Categorical:** Multi-class and binary variables are enforced using enum constraints to restrict output to valid labels.

- **Numerical & Integer:** These are mapped to number and integer types, respectively, with defined range constraints.
- **Datetime:** Temporal data is represented as a string type, governed by a RegEx pattern to ensure strict adherence to the standardized datetime format.

When a column of the original data contains NA values, the corresponding JSON property is combined with the anyOf keyword to a schema with type null, thus allowing the model to optionally generate the null type, which in turn is interpreted as NA in the tabular representation.

##### Enforcing of fuzzy bounds

While providing minimum and maximum constraints for numerical variables ensures data realism, exposing the exact extrema of a clinical dataset can pose significant privacy risks (e.g., identity disclosure of outliers). To mitigate this, we implement fuzzy bounds. Instead of using the raw minimum and maximum values from the original cohort, we enforce boundaries calculated by adding stochastic noise to the observed limits. This ensures the generated values remain within a clinically plausible range without leaking exact boundary information from the source data.

Specifically, the noise is sampled from an exponential distribution defined by $(x_{max} - x_{min})/\varepsilon$, were $\varepsilon$ is a scale parameter representing the noise strength. This sampled value is then added to the observed maximum and subtracted from the observed minimum to define the boundary constraints. For our generations we use a value of $\varepsilon = 10$.

To identify the optimal generative model, we implemented a rigorous selection framework incorporating both fidelity and privacy metrics. This dual-objective analysis allows for the identification of the best performing model, within strictly defined privacy thresholds. While the fuzzy bound strategy structurally prevents the disclosure of exact extrema, the formal assurance of confidentiality is ultimately provided by this empirical selection strategy. The comprehensive methodology for this evaluation is detailed in the main text.

While the fuzzy bound strategy theoretically still allows for the possibility of generating values beyond the observed range, the risk of producing clinically implausible data is significantly mitigated by the exponential decay of the injected noise. Beyond individual variable constraints, a secondary risk involves the generation of plausible values in implausible combinations (e.g., height-weight disparities or incompatible biochemical markers). However, we expect that the LLM can counteract this through its latent domain knowledge acquired during pre-training. By leveraging these internal clinical priors, the model limits the generation of nonsensical feature correlations, further reinforcing the internal consistency and medical validity of the synthetic cohort.

##### Leveraging pre-trained semantic knowledge

To capitalize on the latent clinical knowledge acquired by the LLM during its pre-training phase, we enrich the generation prompts with concise semantic descriptions for each dataset column. These descriptions provide the clinical context necessary for the model to interpret the medical significance of each variable. In turn, this guides the model toward generating values that are not only clinically plausible but also internally consistent across related variables, thereby reducing the risk of generating physiologically nonsensical combinations.

These semantic descriptions also improve the fine-tuning process. By providing a clear linguistic bridge to the model's pre-existing understanding of medical correlations (e.g., the relationship between various lipid variables), the model can focus its learning capacity more efficiently toward capturing the specific statistical patterns and nuances of the target datasets. This approach yields synthetic datasets that maintain a robust general medical foundation while faithfully replicating the intricate correlations found in the original clinical data. A complete list of the column descriptions is provided in the public repository[3].

#### Fine-tuning

For the fine-tuning process, we selected the Qwen3-4B-Base model[4] from the Qwen3 family, initialized with bfloat16 precision. We specifically select the Base versions of the model rather than its instruction-tuned counterpart. Because Base models are not biased toward conversational instruction-following, they generate text based purely on learned probabilistic patterns, making them a more versatile foundation for specialized tasks such as tabular data generation.

##### Training datasets and data partitioning

A significant advantage of the LLM-based approach is its architectural versatility, which allows for the simultaneous fine-tuning of a single generative model on multiple original datasets. This multitasks learning environment enables the model to capture the statistical properties of both cohorts concurrently, potentially "cross-referencing" shared clinical features—such as standard physiological correlations—to develop more robust internal representations. Compared to two separate independent trainings for each source cohort, this multitask approach is particularly advantageous in data-scarce scenarios, as it facilitates a form of knowledge transfer between datasets that can enhance the overall fidelity of the generated synthetic records.

Prior to fine-tuning, we partition the datasets into training (~90%) and test (~10%) sets. To prevent data leakage, we implement consistent splitting: if a patient's record appears in both datasets, it is assigned to the same partition (train or test) in both instances. The test set serves as a strict holdout for final fidelity assessment. Within the training partition, a further 10% is reserved as a validation set to monitor the training loss.

While we monitor the validation loss, we do not rely solely on the minimum validation loss for final model selection. Instead, we save model checkpoints at each validation interval following the minimum loss point. These checkpoints are then subjected to the comprehensive selection procedure detailed in the following section.

To facilitate conditional generation, each training record is split into context columns and completion columns, both serialized into the previously described JSON format. Each training example is structured as a prompt-completion pair:

- **Prompt:** Comprises the JSON representation of the context columns alongside the semantic descriptions of the dataset columns. This segment is excluded from the training loss calculation.
- **Completion:** Comprises the JSON-serialized completion columns and serves as the target for gradient calculation and loss optimization.

The training records from both clinical datasets are shuffled and presented to the model in a randomized sequence.

##### Hyperparameters and configuration

Given the computational intensity of full-parameter fine-tuning, we employ Low-Rank Adaptation (LoRA) to achieve high performance with significantly reduced resource requirements. LoRA freezes the original pre-trained model weights and introduces small, learnable adapter matrices into the transformer layers. In our LoRA configuration we use rank $r = 32$, scaling factor $\alpha = 64$ and dropout rate of 0.05.

The fine-tuning process is implemented using the Hugging Face TRL library[5], integrated with the Unsloth framework[6] to optimize memory efficiency and accelerate training throughput. The models are always trained for exactly 20 epochs, without any early stopping mechanism. Evaluation happens every 100 training steps. We utilize the AdamW optimizer with a fused implementation for enhanced computational performance, maintaining a constant batch size of 16. The learning rate follows a structured schedule:

- **Warmup phase:** The learning rate starts at 0 and increases linearly to a peak of $6\times10^{-5}$ over the first 5% of training steps.
- **Annealing phase:** Following the warmup, a cosine annealing strategy is employed, where the learning rate follows a cosine decay curve, gradually decreasing toward zero.

#### Technical details on model selection

##### Details on the XGBoost discriminator

As outlined in the main text, we use an XGBoost discriminator to compute a distinguishability metric that evaluates synthetic data quality. Because the model is trained to classify records as either real or synthetic, high accuracy reflects a significant deviation from the source distribution, while a low classification rate demonstrates that the synthetic records are statistically indistinguishable from the original ones.

More precisely, the discriminator's AUC-ROC serves as an inverse proxy for fidelity:

- **High AUC-ROC (~1):** Indicates the discriminator can easily distinguish between the two cohorts, signaling low synthetic fidelity.
- **Low AUC-ROC (~0.5):** Indicates the synthetic data is statistically indistinguishable from the real data, signaling high fidelity.

Ideally, the discriminator should be trained and evaluated on a holdout set entirely independent of the generative model's training data. However, given the limited size of our clinical holdout set, we adopt a modified strategy to ensure statistical significance without introducing metric bias. We utilize the generator's training set to train the XGBoost discriminator and reserve the original holdout set exclusively for testing the discriminator's performance. In both phases, we add to the real records an equivalent volume of synthetic data to maintain a balanced class distribution.

To process the tabular data for the XGBoost architecture, we implement the following encoding pipeline:

- **Categorical:** Multi-class and binary variables are transformed using an ordinal encoder, as gradient-boosted decision trees effectively handle ordinal relationships without the dimensionality expansion of one-hot encoding.

- **Numerical & Integer:** Continuous and discrete numerical variables are normalized using a standard scaler.
- **Datetime:** Temporal values are converted to Unix timestamps and subsequently processed with a standard scaler.

#### The computation of the *Privacy Score*

This section details the computation of the *Privacy Score*, the DCR-like metric employed during the model selection process.

As outlined in the main text, establishing a distance-based metric first requires a formal encoding strategy for the dataset records and a corresponding distance function within that encoding space.

We define an attribute-level encoding tailored to the specific data type. For categorical data we use a one-hot encoder, while for integer, numerical, and datetime data (transformed to Unix time) we apply a standard scaler. Once encoded, these attribute-specific numerical vectors are concatenated to form the encoded record representation.

To measure distances between these encoded records, we employ the Euclidean $L_2$ distance, which allows us to leverage optimized algorithms for $k$-NN searches and significantly reduce computational overhead. While alternative metrics exist, we expect our results to be only marginally influenced by the specific choice of distance function.

Given a generative model's training set $R$ and the resulting synthetic dataset $S$, the Privacy Score $P$ is computed as follows:

1. **Perfect synthetic baseline:** Split the training set $R$ into two equal subsets, $R_1$ and $R_2$. Here, $R_2$ serves as a "perfect" synthetic baseline. We then sample a subset of synthetic data $S$ to match the size of $R_1$ and $R_2$.

2. **Distance ratios:** For each record $x$ in $R_1$, we calculate distances to its $k$-th nearest neighbor in $R_1$ (excluding itself), $R_2$ and $S$, denoted as $d_{11}(x)$, $d_{12}(x)$, and $d_{1S}(x)$ respectively. We then compute the ratios $r_{12}(x) = d_{12}/d_{11}$ and $r_{1S}(x) = d_{1S}/d_{11}$.

3. **Statistical comparison:** We construct histograms $h_{12}$ and $h_{1S}$, of $r_{12}(x)$ and $r_{1S}(x)$ for all $x \in R_1$. Intuitively, if synthetic records in $S$ are statistically closer to $R_1$ than the real records in $R_2$, a privacy risk is present. To quantify this, we calculate α, the proportion of records in $h_{1S}$ that fall below the $q$-quantile of $h_{12}$.

4. **Privacy Score:** The Privacy Score is defined as $P = \min(q/\alpha, 1)$.

The Privacy Score $P$ actually represents a family of metrics parameterized by $k$ and the quantile $q$. Because the score probes the left tail of the $h_{1S}$ distribution, lower values of $q$ generally yield a more sensitive measure of the privacy risk; however, this sensitivity comes at the cost of increased variance due to the smaller sample size involved in the calculation. As defined, $P$ ranges from $q$ (indicating high residual risk) to 1 (indicating low risk). In our analysis, we use $k = 1$ and $q = 0.1$.

#### Anonymeter attacks

The Anonymeter framework defines three distinct attack vectors, each targeting a specific dimension of privacy risk:

1. **Singling out:** This attack involves generating a set of logical predicates from the synthetic dataset to determine if they uniquely identify a single record within the target (training or control) data. Two types of predicates are available:
    - **Univariate predicates:** These check for unique values within individual columns. For numerical attributes, additional predicates are generated based on observed extrema. However, we observe that univariate checks for numerical columns yield inconsistent results in our context—particularly with our implementation of classical anonymization. Consequently, we exclude univariate predicates from our final assessment.
    - **Multivariate predicates:** These are constructed as logical AND combinations of univariate predicates derived from randomly selected records and attributes. Our analysis of singling-out risk focuses exclusively on these multivariate combinations.

- **Linkability:** The linkability attack models the ability of an adversary to re-identify individuals by connecting disparate data fragments. The attacker possesses a set of real target records $T$, bifurcated into two subsets, $T_A$ and $T_B$, containing complementary attribute sets $A$ and $B$, respectively. By cross-referencing the $k$ nearest synthetic neighbors for each record in $T_A$ and $T_B$, the attacker attempts to link the fragments belonging to the same individual. For this evaluation, we utilize the default parameter of $k = 1$ and perform a randomized split of attributes to define the $A$ and $B$ subspaces. Notably, MIAs are sometimes viewed as a way to model linkability.

- **Inference:** In an inference attack, the adversary possesses a partial set of attributes for certain target original records and attempts to predict the remaining "secret" attributes. The inference "guess" is derived from the closest synthetic record within the known attribute subspace. A guess is considered successful if it exactly matches the categorical secret attributes or falls within a specified tolerance δ, for numerical values. In our configuration, we maintain the default tolerance of δ and select a single target secret attribute.

#### Classical anonymization

To contextualize the performance of our LLM-generated synthetic data, we benchmark our results against datasets processed through traditional anonymization techniques. For this purpose, we utilize the aindo.anonymize Python library[7], which implements several column-wise anonymization techniques. Specifically, we use:

- PerturbationCategorical for categorical and boolean variables. With this perturbation mechanism each record element is replaced with probability α, by a value randomly sampled from the set of all possible categories. The sampling logic is governed by the sampling_mode parameter:
    - **Uniform mode:** Each category within the attribute's domain is equally likely to be selected.

- **Weighted mode:** The replacement values are sampled according to the original frequency distribution of the categories in the source data.

- PerturbationNumerical for numerical, integer and datetime variables (transformed to Unix time). It incorporates random noise into the original values, governed by a mixing parameter α. The noise injection follows one of two methodologies—according to the sampling_mode parameter:
  - **Uniform mode:** Stochastic noise, sampled from a uniform distribution within the attribute's observed range $[x_{min}, x_{max}]$, is added to the original data points.
  - **Weighted mode:** To preserve the underlying one-way marginal distribution of each column, the original values are first mapped to a standard Gaussian distribution using a quantile transformer. Gaussian noise is then injected within this transformed space before the data is back-transformed into its original scale.

In our evaluation, we utilize the weighted sampling mode for all perturbations. We systematically vary the noise intensity parameter α, across a range from 0.1 to 0.9. This produces a family of anonymized datasets with a gradient of noise levels. As α increases, we anticipate a characteristic trade-off: a strengthening of privacy guarantees at the expense of a corresponding decrease in data fidelity. Notice that we use the same value of α for both categorical and numerical perturbations, even if in principle the two parameters are not related.

Unlike synthetic data generation, which produces entirely new records, classical anonymization maintains a strict one-to-one correspondence between the original and perturbed records. To ensure a valid comparison using the fidelity and privacy metrics previously discussed, we maintain this correspondence across all data partitions. Specifically, whenever the original dataset is bifurcated into training and test sets, the anonymized data is partitioned consistently. For example, to evaluate fidelity via the XGBoost discriminator, the training set is constructed from the union of a subset of the original records and their directly corresponding anonymized versions. The remaining original records and their respective anonymized counterparts form the holdout set for the discriminator's performance assessment. This consistent partitioning ensures that the discriminator evaluates the distributional shift caused by anonymization without being confounded by the accidental inclusion of identical or closely related records across the train-test split.

The classical anonymization techniques employed in this study belong to the randomization family; however, they represent only one subset of available de-identification strategies. Another frequently used randomization method is swapping, which rearranges data by randomly exchanging values between records. A different family is generalization, which includes techniques such as binning, where numerical values are aggregated into discrete intervals to ensure multiple individuals share identical attributes. Additionally, suppression may be applied to mask rare categorical values or numerical extrema by replacing them with a constant. These methods are frequently used to achieve $k$-anonymity, which ensures that each record is indistinguishable from at least $k - 1$ others based on a set of quasi-identifiers. Further refinements include $l$-diversity, which requires at least $l$ well-represented values for each sensitive attribute within each anonymized group, and $t$-closeness, which demands the distribution of sensitive attributes within each anonymized group is $t$-close to the global distribution.

Our selection of randomization techniques is specifically dictated by the necessity of a valid comparison; the anonymized datasets must remain evaluable under the same fidelity and privacy metrics applied to the real and synthetic data. By utilizing these specific techniques, the anonymized data maintains the same structure as the original dataset, preserving the continuous and categorical formats required for a direct comparison. In contrast, alternative methods may have necessitated a different evaluation framework. For instance, distinguishability analysis via an XGBoost discriminator would yield trivial results if applied to binned or suppressed data, as the model would identify structural artifacts rather than statistical distributions. Furthermore, unlike discrete de-identification methods, our implementation of randomization allows for the continuous calibration of noise intensity through the $\alpha$ parameter, enabling a granular exploration of the privacy-utility trade-off. Ultimately, we utilize these anonymized benchmarks as a representative baseline and do not anticipate that alternative classical methods would yield a significantly better privacy-utility profile.

### References


[1] https://docs.vllm.ai/en/stable/

[2] https://github.com/guidance-ai/llguidance

[3] https://github.com/aindo-com/monzino-sams-ldl

[4] https://huggingface.co/Qwen/Qwen3-4B-Base

[5] https://huggingface.co/docs/trl/index

[6] https://unsloth.ai/

[7] https://docs.anonymize.aindo.com/latest/

### Supplemental tables

*Supplemental Table 1. Mapping between column types and JSON representation.*

| **Column type** | **JSON type** | **Validation keywords** |
|---|---|---|
| Categorical | - | enum |
| Numeric | number | minimum, maximum (fuzzy) |
| Integer | integer | minimum, maximum (fuzzy) |
| Datetime | string | pattern |
